# Genetic Risk Factors for Severe and Fatigue Dominant Long COVID and Commonalities with ME/CFS Identified by Combinatorial Analysis

**DOI:** 10.1101/2023.07.13.23292611

**Authors:** Krystyna Taylor, Matthew Pearson, Sayoni Das, Jason Sardell, Karolina Chocian, Steve Gardners

**Affiliations:** PrecisionLife Ltd, Unit 8b Bankside, Hanborough Business Park, OX29 8LJ, UK

**Keywords:** Long COVID, post-acute sequelae of COVID-19, PASC, post-Covid, post-acute COVID syndrome, POTS, ME/CFS, patient stratification, combinatorial analytics

## Abstract

**Background:** Long COVID is a debilitating chronic condition that has affected over 100 million people globally. It is characterized by a diverse array of symptoms, including fatigue, cognitive dysfunction and respiratory problems. Studies have so far largely failed to identify genetic associations, the mechanisms behind the disease, or any common pathophysiology with other conditions such as ME/CFS that present with similar symptoms.

**Methods:** We used a combinatorial analysis approach to identify combinations of genetic variants significantly associated with the development of long COVID and to examine the biological mechanisms underpinning its various symptoms. We compared two subpopulations of long COVID patients from Sano Genetics’ Long COVID GOLD study cohort, focusing on patients with severe or fatigue dominant phenotypes. We evaluated the genetic signatures previously identified in an ME/CFS population against this long COVID population to understand similarities with other fatigue disorders that may be triggered by a prior viral infection. Finally, we also compared the output of this long COVID analysis against known genetic associations in other chronic diseases, including a range of metabolic and neurological disorders, to understand the overlap of pathophysiological mechanisms.

**Results:** Combinatorial analysis identified 73 genes that were highly associated with at least one of the long COVID populations included in this analysis. Of these, 9 genes have prior associations with acute COVID-19, and 14 were differentially expressed in a transcriptomic analysis of long COVID patients. A pathway enrichment analysis revealed that the biological pathways most significantly associated with the 73 long COVID genes were mainly aligned with neurological and cardiometabolic diseases.

Expanded genotype analysis suggests that specific *SNX9* genotypes are a significant contributor to the risk of or protection against severe long COVID infection, but that the gene-disease relationship is context dependent and mediated by interactions with *KLF15* and *RYR3*.

Comparison of the genes uniquely associated with the Severe and Fatigue Dominant long COVID patients revealed significant differences between the pathways enriched in each subgroup. The genes unique to Severe long COVID patients were associated with immune pathways such as myeloid differentiation and macrophage foam cells. Genes unique to the Fatigue Dominant subgroup were enriched in metabolic pathways such as MAPK/JNK signaling. We also identified overlap in the genes associated with Fatigue Dominant long COVID and ME/CFS, including several involved in circadian rhythm regulation and insulin regulation. Overall, 39 SNPs associated in this study with long COVID can be linked to 9 genes identified in a recent combinatorial analysis of ME/CFS patient from UK Biobank.

Among the 73 genes associated with long COVID, 42 are potentially tractable for novel drug discovery approaches, with 13 of these already targeted by drugs in clinical development pipelines. From this analysis for example, we identified TLR4 antagonists as repurposing candidates with potential to protect against long term cognitive impairment pathology caused by SARS-CoV-2. We are currently evaluating the repurposing potential of these drug targets for use in treating long COVID and/or ME/CFS.

**Conclusion:** This study demonstrates the power of combinatorial analytics for stratifying heterogeneous populations in complex diseases that do not have simple monogenic etiologies. These results build upon the genetic findings from combinatorial analyses of severe acute COVID-19 patients and an ME/CFS population and we expect that access to additional independent, larger patient datasets will further improve the disease insights and validate potential treatment options in long COVID.

## Introduction

Post COVID-19 condition (or long COVID) is a debilitating syndrome that the World Health Organization (WHO) estimates affects up to 20% of people infected by SARS-CoV-2^1^. Other more recent studies put the prevalence of long-term symptoms (over 3 months post-infection) in COVID-19 patients even higher^2^, with all estimates implying that over 100 million patients have been affected by the condition globally^3^. Even though symptoms decline for most patients over time, some patients still experienced symptoms such as post-exertional malaise or postural tachycardia syndrome (POTS)^4^ up to two years after infection^5^, and the long-term health consequences of long COVID remain unknown, with suggestions of a doubling of the risk of developing cardiovascular issues^6^.

Reports indicate an extensive array of symptoms associated with long COVID^7^, with the most common being fatigue and post-exertional malaise (PEM)^8^, cognitive dysfunction^9^, mood disturbances^10^ and respiratory problems^11^. However, establishing a precise diagnosis for either of these diseases has proved challenging, in large part due to the complexity and diversity of their clinical presentation and their effects across multiple organ systems. In an attempt to provide some definitive metrics, a recent study developed a data-driven scoring framework for diagnosing long COVID based on the available symptom data^12^.

Although many studies have investigated the genetic risks underlying long COVID, only one genome-wide association study (GWAS) has identified a single risk locus around the lead variant in *FOXP4*^13, 14^. Studies that used combinatorial analytical approaches to delineate genetic risk factors in similarly heterogenous populations have demonstrated more success, for example in severe COVID-19^15^ and ME/CFS^16^.

Combinatorial analytics approaches identify combinations of features that together (rather than individually) are associated with the disease phenotype^17^. They capture the non-linear effects of interactions between multiple genes (and exogenous factors if available). These signals are distinct from and complementary to the monogenic, linear additive associations of single SNPs found by GWAS. In complex (multifactorial and heterogenous) diseases these non-linear combinatorial signals are significantly more important in understanding disease biology than in relatively monogenic disorders such as many cancers and rare genetic disorders^18, 19^.

In this study we used combinatorial analytics to identify disease risk signatures (combinations of genetic variants significantly associated with the development of long COVID) and explored the biological mechanisms with which they are involved. We investigated subpopulations of long COVID patients who had experienced either severe disease or a fatigue dominant phenotype, to compare the underlying genes and pathways that explain some of the heterogenous manifestations of the disease.

We also compared the output of this study against our previous ME/CFS analysis^16^ to understand similarities in post-viral fatigue and other phenotypes experienced by subsets of long COVID patients. Finally, we compared the pathways that were significantly enriched in this genetic analysis of long COVID against known genetic associations in other chronic diseases that are predominantly autoimmune, neurological and/or metabolic in nature, to evaluate any common pathophysiological mechanisms that might be shared by long COVID.

## Methods

### Sano Genetics GOLD Study Dataset

Genotypic and phenotypic data for both cases and controls included in this study were generated from Sano Genetics’ Long COVID GOLD study^20^. Eligible participants (n = 1,996), recruited between 2020 and 2022, provided saliva samples for an at-home Sano DNA Test (evaluated via Illumina Global Screening Array with Multi-disease drop-in panel) and completed a questionnaire hosted on the Sano Genetics platform detailing their acute COVID-19 and long COVID symptoms (if experienced), as well as basic demographic data and other chronic health conditions (see Supplementary File 1).

### Symptom Based Score for Long COVID Severity

Given the heterogeneity of post-COVID symptoms reported by the GOLD study and other previous studies, we developed a data-driven scoring method to characterize the severity of self-reported symptoms. We analyzed participant reported scores for each available long COVID symptom experienced pre- and post-acute COVID-19, including breathlessness, fatigue, degree of muscle pain and change in mental health (see Supplementary Table 1 for more details). A ‘Total Change’ score was generated for each patient from the sum of the reported differences across symptoms pre- and post-COVID.

### Cohort Characteristics

At the time of analysis, a total of 1,829 individuals in the GOLD study had a self-reported COVID-19 diagnosis. This COVID-19 cohort had a median age of 50 years [interquartile range (IQR) = 40 - 60] and median COVID-19 recovery time of 169 days [IQR = 14 - 507.5] (Table 1). It consisted of 61.1% females and 92.6% self-reported their ethnicity as ‘White’. The most prevalent self-reported comorbidities (prior to or after COVID-19) in the cohort were anxiety or panic attacks (30.0%), depression (26.2%), asthma (25.5%), eczema (18.6%) and migraines (17.4%).

**Table 1:** Characteristics of the two long COVID cohorts derived from the GOLD study dataset. Data for fields marked with asterisk (*) were not available for all individuals. Comorbidities marked with ^†^ were consistently over-represented in cases compared to controls in all cohorts.

| | Severe Long COVID<br>$n = 1,323$ | | Fatigue Dominant Long COVID<br>$n = 1,386$ | |
| --- | --- | --- | --- | --- |
| | Cases<br>($n=459$) | Controls<br>($n=864$) | Cases<br>($n=477$) | Controls<br>($n=909$) |
| **Age [Median (IQR)]** | 45 (37-54) | 54 (41-64) | 45 (37-54) | 54 (41-63) |
| **Sex [$n$ (%)] \*** | M: 129 (28.1)<br>F: 329 (71.7) | M: 402 (46.5)<br>F: 462 (53.5) | M: 121 (25.4)<br>F: 355 (74.4) | M: 429 (47.2)<br>F: 480 (52.8) |
| **Self-reported Ethnicity [$n$ (%)] \***<br>Wh = White<br>As = Asian<br>Mx = Mixed<br>Bl = Black<br>Ot = Other<br>No = None | Wh: 419 (91.3)<br>Mx: 19 (4.1)<br>As: 12 (2.6)<br>Ot: 4 (0.9)<br>Bl: 2 (0.4)<br>No: 2 (0.4) | Wh: 781 (90.4)<br>As: 33 (3.8)<br>Mx: 22 (2.5)<br>Bl: 12 (1.4)<br>Ot: 12 (1.4)<br>No: 1 (0.1) | Wh: 436 (91.4)<br>Mx: 17 (3.6)<br>As: 14 (2.9)<br>Bl: 4 (0.8)<br>Ot: 3 (0.6)<br>No: 2 (0.4) | Wh: 825 (90.1)<br>As: 33 (3.6)<br>Mx: 23 (2.5)<br>Ot: 13 (1.4)<br>Bl: 11 (1.2)<br>No: 1 (0.1) |
| **Recovery time in days (Median [IQR] )** | 479<br>(247-572) | 18<br>(8-122) | 484<br>(256-573) | 18<br>(8-111) |
| **COVID-19 related hospitalization [n (%)]** | 60 (13.1) | 26 (3.0) | 66 (13.8) | 26 (2.9) |
| **Reported problems after COVID-19 related hospital discharge [n (%)]** | 60 (13.1) | 20 (2.3) | 66 (13.8) | 19 (2.1) |
| **Hospitalized [n (%)]** | 46 (10.0) | 7 (0.8) | 50 (10.5) | 8 (0.9) |
| **Co-morbidities (pre-existing or post-COVID-19) [n (%)]** |  |  |  |  |
| - **Asthma<sup>†</sup>** | 108 (23.5) | 121 (14.0) | 110 (23.1) | 133 (14.6) |
| - **Alzheimer's disease** | 0 (0.0) | 1 (0.1) | 0 (0.0) | 1 (0.1) |
| - **Coronary artery disease** | 2 (0.4) | 13 (1.5) | 3 (0.6) | 13 (1.4) |
| - **Chronic fatigue syndrome<sup>†</sup>** | 36 (7.8) | 6 (0.7) | 40 (8.4) | 5 (0.6) |
| - **Diabetes Type 1** | 1 (0.2) | 9 (1.0) | 1 (0.2) | 10 (1.1) |
| - **Diabetes Type 2** | 15 (3.3) | 33 (3.8) | 13 (2.7) | 33 (3.6) |
| - **Heart attack** | 5 (1.1) | 7 (0.8) | 3 (0.6) | 7 (0.8) |
| - **Irritable bowel syndrome<sup>†</sup>** | 65 (14.2) | 41 (4.7) | 74 (15.5) | 47 (5.2) |
| - **Kidney disease<sup>†</sup>** | 3 (0.7) | 2 (0.2) | 3 (0.6) | 2 (0.2) |
| - **Liver disease<sup>†</sup>** | 9 (2.0) | 6 (0.7) | 8 (1.7) | 7 (0.8) |

**Table 2:** Summary of PrecisionLife combinatorial analysis results on Severe and Fatigue Dominant long COVID cohorts generated from the GOLD study.

|  | Severe Cohort | Fatigue Dominant Cohort |
| --- | --- | --- |
| Total disease signatures (*n*) | 1,188 | 1,435 |
| Disease signatures by number of component SNPs (*n*) | 0 , 2 , 88 , 322 , 776 | 0 , 25 , 191 , 225 , 994 |
| Odds ratio of disease signatures (relative to mean odds, Median [Q1-Q3]) | 77.9 [19.5 - 80.0] | 22.5 [9.4 - 23.6] |
| RF scored "critical" SNPs (*n*) | 86 | 84 |
| RF scored genes (*n*) | 43 | 36<br>(35 after ancestry confounder check) |
| Cases containing at least one disease signature (%) | 100% | 100% |

Of those confirmed to have had COVID-19, 1,345 (73.5%) reported fatigue symptoms, 1,135 (62.1%) reported symptoms linked to concentration, 1,124 (61.5%) reported short-term memory symptoms and 714 (39.0%) reported breathlessness. The median ‘Total Change’ symptom score for the cohort was 15 [IQR = 2 - 35] (Supplementary Figure 1).

In the dataset, 1,489 (81.4%) individuals provided free-text responses on other symptoms that they experienced since their illness that were not covered elsewhere in the questionnaire. The most frequently reported symptoms included loss of smell, headache, pain, tinnitus, loss of taste, dizziness, insomnia and postural tachycardia syndrome (POTS) (see Supplementary Table 2 and Supplementary Figure 2). Following COVID infection, 353 (19.3%) individuals reported reducing their working hours while 359 (19.6%) people discontinued working altogether post-illness.

### Long COVID Cohorts

We defined two long COVID case populations from the GOLD study based on self-reported symptom changes three months post COVID-19 – ‘Severe’ long-haulers who reported the greatest variety and severity of symptoms and ‘Fatigue Dominant’ cases who reported predominantly fatigue-associated long COVID symptoms.

The World Health Organization defines long COVID patients as those experiencing one or more symptoms post initial COVID-19 infection. However, the cohort in the GOLD study that met these criteria displayed a great range in the severity and length of self-reported symptoms experienced post COVID-19. Instead, we aimed to focus on the more ‘severe’ long haulers who reported the greatest degree of symptoms experienced as these are likely to be the patients experiencing long COVID symptoms that do not diminish over time without pharmaceutical intervention.

The Fatigue Dominant’ cohort was chosen primarily due to their phenotypic similarity with ME/CFS, allowing us to explore potential commonalities between the diseases based on our previously published combinatorial analysis for ME/CFS^16^. The number and overlap in cases and controls included in the two datasets are included in Supplementary Figure 3.

#### Severe Long COVID Cohort

The Severe long COVID cohort (*n* = 1,323 where cases = 459 and controls = 864) was selected using the difference in scores reported pre- and post-acute COVID-19 for three long COVID symptom groups – namely, respiratory, fatigue and mental health. Severe cases were defined as those with a ‘Total Change’ score for these symptoms greater than or equal to the upper quartile of the distribution. The controls in this study were defined as samples with a ‘Total Change’ score greater than or equal to 0 but below the median of the distribution.

#### Fatigue Dominant Long COVID Cohort

The Fatigue Dominant cohort (*n* = 1,386 where cases = 477 and controls = 909) was selected using only a subset of symptoms relating to fatigue in the scores (‘Fatigue Change’) reported for pre- and post-acute COVID-19 symptoms (see Supplementary Table 1). The controls in this study were defined as samples with a ‘Fatigue Change’ score greater than or equal to 0 but below the median of the distribution. The characteristics of the two cohorts are described in Table 1, Figure 1 and Supplementary Figure 4.

**Figure 1:**
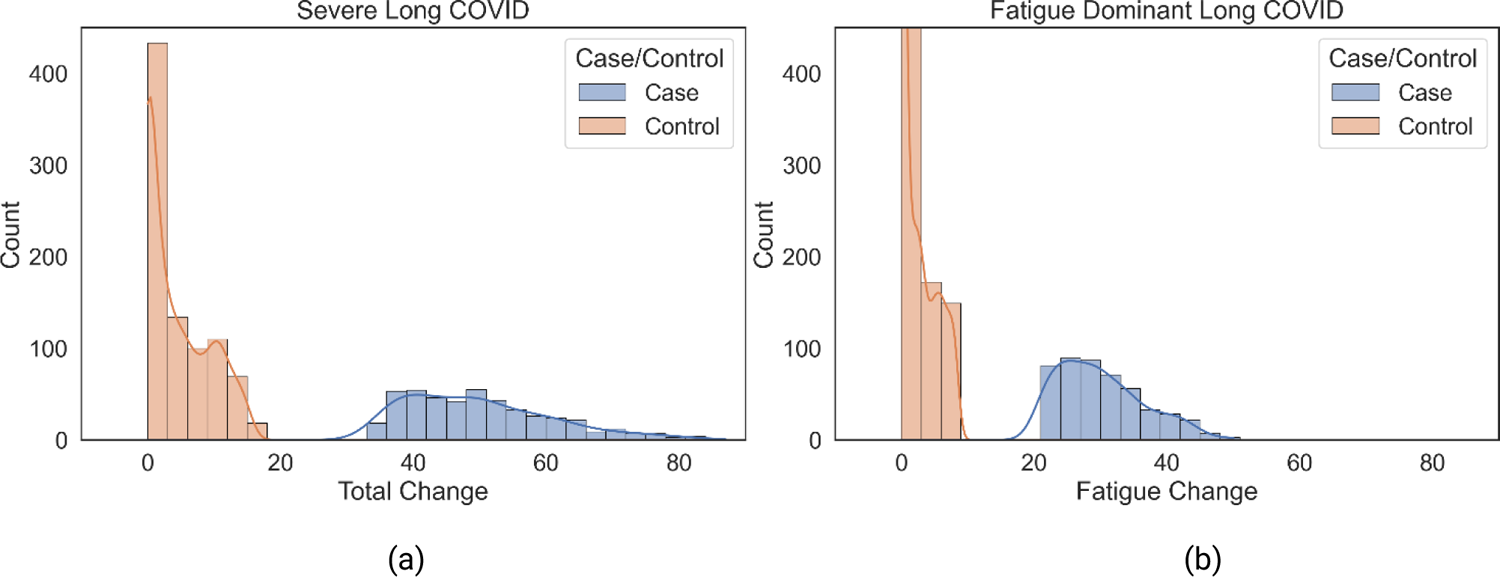
Distribution of the (a) ‘Total Change’ score for cases and controls in the Severe long COVID and (b) ‘Fatigue Change’ (part of ‘Total Change’ score) score in the Fatigue Dominant long COVID cohorts.

### Dataset QC

The two case-control datasets underwent a series of quality control (QC) procedures before they were analyzed using the PrecisionLife platform.

Standard variant-level and sample-level QC procedures were applied to the dataset (comprising of 696,382 SNPs) as described in the Genotype Quality Control section in Supplementary Information. Due to the small sample size of the two long COVID cohorts, the genotype data was filtered to exclude SNPs with minor allele frequency (MAF) < 5%. Very low frequency SNPs were removed as significant combinations involving rare variants are especially infrequent. This filter also increases the statistical power of combinatorial analysis to detect genotype-disease associations by reducing the amount of false discovery rate (FDR) correction required when testing multiple SNP-genotype combinations. Following QC, the Severe dataset comprised of 283,478 SNPs and the Fatigue Dominant dataset contained 283,444 SNPs.

### Combinatorial Analytics using the PrecisionLife Platform

The PrecisionLife combinatorial analysis platform enables hypothesis-free identification of high-order combinatorial features (known as disease signatures), which may include multiple SNP genotypes and/or other multi-modal features in combination. These disease signatures capture both the linear and non-linear effects of genetic and molecular interaction networks and enable the identification of associations including those that are only relevant to a subgroup of patients. We have previously validated this analytical approach across a variety of complex chronic diseases where it has identified more associations with increased explanation of observed disease variance and reproducibility than comparable GWAS studies^15–17^.

In the combinatorial analytics approach, disease signatures are identified and statistically validated in ‘layers’ of increasing combinatorial complexity, i.e., singletons, pairs, triplets etc. (also known as combinatorial order). Each disease signature is validated multiple times using several statistical tests at each stage of the process to avoid false positives. A more detailed description of the mining and validation stages is given in our previous ME/CFS study^16^.

We applied the PrecisionLife platform to both long COVID case-control datasets in a hypothesis-free manner to identify combinations of SNP genotypes that are strongly associated with the development of long COVID symptoms when they co-occur in the same patient. The method prioritizes SNP genotype combinations that have high odds ratios, low *p*-values (*p* < 0.05) and high prevalence (>5%) in long COVID cases. A permutation-based approach was used to compare the observed properties of the most highly associated SNP-genotype combinations to the null distribution for randomized datasets^21^, with *p*-value cut-offs based on a specified threshold (Benjamini-Hochberg FDR of 0.05) after multiple testing correction. Combinations passing these tests were reported as validated long COVID disease signatures. Finally, a merged network (disease architecture) view is generated by clustering all validated disease signatures based on their co-occurrence in patients in the dataset.

SNPs found in multiple disease signatures often form the central hub of the disease architecture (see Figure 2). These are termed ‘critical SNPs’ if the corresponding networks pass a further permutation-based statistical test. Potential critical SNPs are scored using a Random Forest (RF) algorithm with a 5-fold cross-validation framework to assess the accuracy with which they predict the case-control split in the dataset.

**Figure 2:**
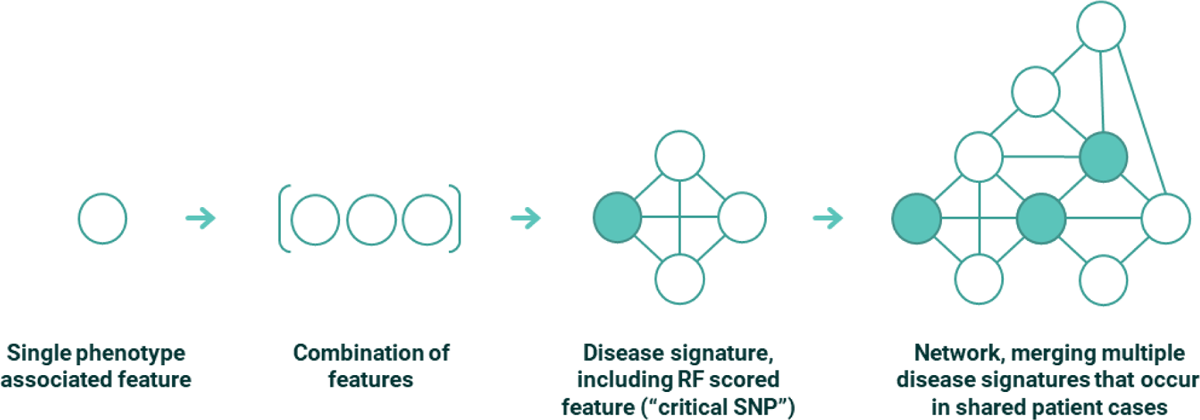
Conceptual representation of features, combinations and disease signatures that form part of PrecisionLife’s combinatorial analytics methodology. In the case of the long COVID study all features were SNP genotypes, but other feature types, e.g., a patient’s expression level of a specific protein, medication history or clinical features such as their eosinophil level, can also be used, independently from or in combination with the genotype data.

A cascade mapping process was used to map all the critical SNPs identified in the validated disease signatures to the human reference genome (GRCh38)^22^. SNPs identified in the coding region of a gene (or genes) were mapped directly to this gene and any remaining SNPs within 2kb upstream or 0.5kb downstream were mapped to the nearest gene(s). Due to the uncertainty about the wide range of cells and tissues that have been implicated in long COVID etiology^7^, genes assigned by either expression quantitative trait loci (eQTLs) or chromatin interaction (Hi-C) data were not specifically prioritized for further analysis (as they would likely be in other indications) to avoid capturing any spurious associations from non-trait-related tissues or cells. Genes that could additionally be mapped using only eQTL or Hi-C data from the critical SNPs were observed and reported in Supplementary File 2, although these were not further evaluated.

Finally, a semantic knowledge graph, including data from over 50 public data sources (see Supplementary Table 3), was used to annotate the SNPs and genes, including data on prior genetic associations to disease, chromosomal location, tissue expression profiles, splice variants, mouse phenotypes, protein function/structure, known active chemistry and any pre-existing scientific literature or clinical trials among other attributes. This allows us to generate evidence-backed mechanism of action hypotheses as to each genetic variant’s potential impact on a patient’s long COVID phenotype.

### Ancestry Analysis

Ancestry inference for the samples in the GOLD study was performed using GRAF-pop^23^. To maximize the number of samples included in each case-control dataset, samples of all ancestries were included in the analysis. Since ancestry-specific analyses could not be performed due to limited samples in each cohort, we performed a logistic regression analysis to control for confounding effects of population structure. Any disease signatures that were no longer significantly associated with case-control status (*p* < 0.05 with Bonferroni FDR correction) in a logistic regression that also includes a binary ancestry variable for white-European/other ancestry were considered false positives and removed from further analysis.

### Assessing Causality with Expanded Genotypes Analysis

The disease signatures output by the PrecisionLife platform represent combinations of SNP genotypes that are significantly enriched in cases relative to controls. Expanded genotypes analysis (“EGA”) tests how the genotype of a critical SNP from the disease signature affects the odds of disease when the genotypes of all interacting SNPs are held constant.

For each disease signature, we first assign patients to one of the possible combinations of the component SNP genotypes (the “expanded genotype signatures”). In the example illustrated in Figure 3, the validated disease signature is comprised of two SNPs, each in one of 3 states (0, 1 and 2), which can generate 9 (3^2^) expanded genotype signatures. For combinations of 3, 4, and 5 SNPs, the number of expanded genotypes signatures is 27, 81, and 243 respectively. We then calculate the disease odds for patients with each expanded genotype signature.

**Figure 3:**
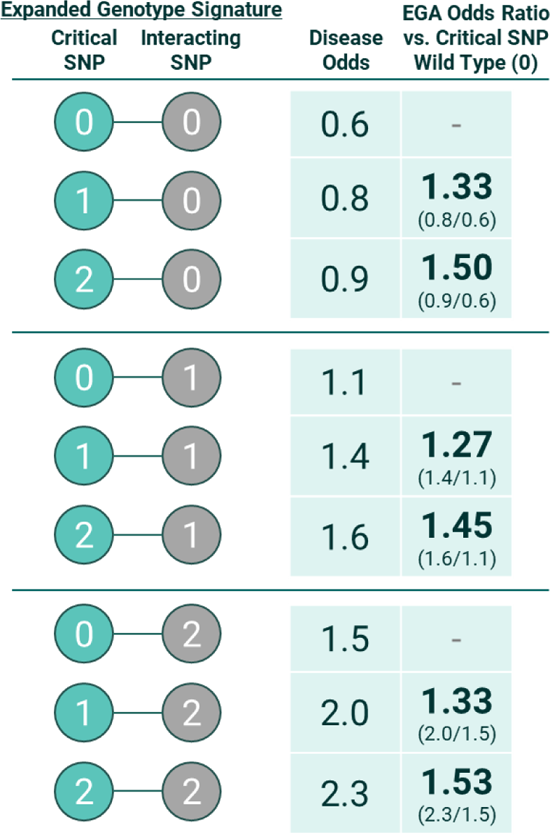
Hypothetical example of an expanded genotypes analysis for a disease signature comprised of two SNPs. After controlling for the confounding effects of the interacting SNP genotype, patients with one or two copies of the critical SNP minor allele (genotypes “1” and “2”) have consistently elevated odds of disease relative to patients with the wild type genotype (“0”) at the critical SNP.

For a given critical SNP of interest, we identify sets of expanded genotype signatures that share the same genotypes for all interacting SNPs (the blocks separated by the horizontal lines in Figure 3). We calculate the “EGA odds ratio” by dividing the disease odds ratio for an expanded genotype signature with a copy of the critical SNP minor allele by the disease odds ratio for the matching expanded genotype signature with the critical SNP homozygous wild type genotype.

Due to the small number of patients associated with individual expanded genotype signatures, we may have insufficient statistical power to directly test whether the EGA odds ratios are significantly different from zero. Instead, the primary aim of the EGA is to test whether the observed directionality of the relationship between the critical SNP minor allele and disease phenotype is consistent across all or most expanded genotype signatures. If the critical SNP genotype does not affect disease, then we expect the minor allele genotype will be randomly associated with increased odds of disease for some expanded genotype signatures and decreased odds of disease for others, with no consistent biological pattern.

In the hypothetical example shown in Figure 3, the EGA reveals that the critical SNP minor allele is consistently associated with elevated disease risk after controlling for the genotype of the interacting SNP. This pattern holds even though patients with the critical SNP minor allele have below average odds of disease when they also possess the wild type genotype at the interacting SNP. By controlling for the confounding effects of the interacting SNP, EGA allows us to gain a better understanding of the relationship between the critical SNP and disease.

Each disease signature was assigned to one of the following seven categories based on the broad patterns observed from the EGA: universally causative, universally protective, SNP-specific causative, SNP-specific protective, combination-specific causative, combination-specific protective, or ambiguous. Definitions of each category are provided in Supplementary Table 5. Across these categories, the designation of “Causative” and “Protective” do not necessarily guarantee that the specific critical SNP identified in the analysis directly affects disease risk. The critical SNP could potentially be a neutral marker that is in strong linkage disequilibrium with the true biological variant.

We excluded all expanded genotype signatures which occurred in fewer than 15 patients from the EGA. Likewise, we did not consider disease signatures comprised of 4 or 5 SNPs due to the limited statistical power provided by the size of the available datasets. There are 81 possible expanded genotype signatures for a combination of 4 SNPs, which corresponds to only 17 patients per expanded genotype signature. More problematically, there are 243 possible expanded genotype signatures for a combination of 5 SNPs, which corresponds to fewer than 6 patients per expanded genotype signature. The stochastic noise associated with such small sample sizes make it very difficult to identify broad patterns across the full set of expanded genotype pairs.

### Phenotype Enrichment Analysis

The available clinical data from the questionnaire was used to evaluate the long COVID patient profiles associated with each of the disease signatures generated by the analysis. We calculated the statistical significance of the association of a particular phenotype with a set of long COVID cases with shared genetic variants when compared against the rest of the case population. The two proportions Z-test was used for categorical variables, such as severity of acute COVID-19 and comorbidities, and Mann-Whitney U^24^ for any continuous variables, such as participant reported scores that reflect change in symptoms pre- and post-COVID-19. Statistical associations were corrected for multiple testing using Benjamini-Hochberg method.

### Overlap Analysis (“Seeded” Approach)

We evaluated the genetic overlap between the Severe and Fatigue Dominant cohorts by taking the SNPs identified in the hypothesis-free analysis for one dataset (seed SNPs) and testing whether any combinations involving them are also significantly associated with disease risk in the second dataset when analyzed by the PrecisionLife platform (see section ‘Combinatorial Analytics using the PrecisionLife Platform’).

This hypothesis-driven or ‘seeded’ approach was performed in addition to a direct gene overlap analysis between the two cohorts. This approach mitigates the effects of stochastic differences in dataset composition when defining the combinatorial search space explored in our analyses. The number of possible SNP-genotype combinations is so extensive that it is impossible to sample the entirety of the space. This implies that true associations may remain unreported because they were not tested when the dataset was analyzed using the hypothesis-free approach.

We also employed this technique when evaluating the overlap between the genes identified in our analysis of the UK Biobank ME/CFS population and the two long COVID cohorts generated from the GOLD study. Due to the low SNP overlap (*n*=42,500) between the arrays used to genotype the ME/CFS and long COVID datasets, we performed a seeded analysis using 383 SNPs in the Severe and Fatigue GOLD dataset that were within 10kb up or downstream of the original 14 ME/CFS genes.

### Cross Disease Analysis

Cross disease analysis can provide insights into potential drug repurposing opportunities or development of common therapies. We compared the genes that were significantly associated with Severe and Fatigue Dominant long COVID against a variety of other chronic diseases to identify shared pathophysiological mechanisms. These diseases included neurodegenerative, mental and behavioural disorders, cardiovascular, gastrointestinal, autoimmune and metabolic diseases (see Supplementary Tables 8 and 9). Disease-associated genes identified for each indication group are those with known genetic links reported in OpenTargets^25^ (v 23.02, February 2023 release). Only genes with strong target-disease genetic association scores (>0.9 out of 1.0) have been used in this analysis for each indication group.

Enrichment analysis was performed using the g:Profiler tool^26^ to determine pathways and biological processes that are significantly associated with the disease-associated genes for each indication group (*p* < 0.05, *p*-value correction for multiple testing using Benjamini-Hochberg). This allows us to explore up/downstream of individual gene targets to identify biological processes that are impacted across diseases.

## Results

### GWAS Analysis

We evaluated the significance of individual genetic variants associated with the two long COVID datasets (Severe and Fatigue Dominant) using a standard GWAS analysis with PLINK^27^. As can be observed from the two Manhattan plots (Supplementary Figure 5), no SNP from either of the two cohorts reached the genome-wide significance threshold (*p* < 5×10^−8^).

### Hypothesis Free Combinatorial Analysis

Using the PrecisionLife combinatorial analysis platform, we identified 86 disease associated critical SNPs for the Severe cohort and 84 for the Fatigue Dominant cohort, mapping to 43 and 36 genes respectively. A total of 74 unique genes were associated with at least one of the long COVID cohorts, including 5 genes which were identified in both the Severe and Fatigue Dominant cohorts.

The disease signatures associated with each cohort were all combinations of 2 or more SNP genotypes, i.e., they were all combinatorial signals, predominantly involving combinations of 3-5 SNPs, that could not have been identified using GWAS (Figure 4). An example of one of the disease signatures identified in the analysis of the Severe long COVID cohort is shown in Table 3. None of the SNPs identified in disease signatures were observed to be in linkage disequilibrium (LD) with each other.

**Figure 4:**
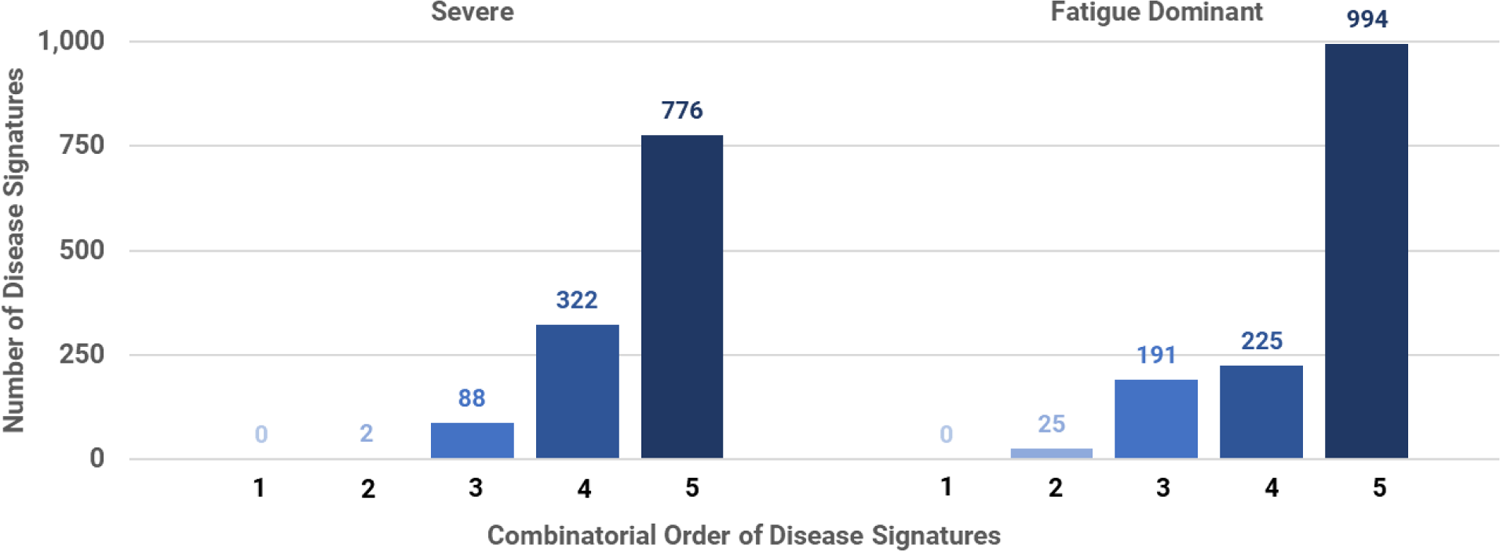
Distribution of combinatorial order (i.e., number of component SNPs) for the validated combinatorial disease signatures from the Severe and Fatigue Dominant long COVID cohorts.

**Table 3:**
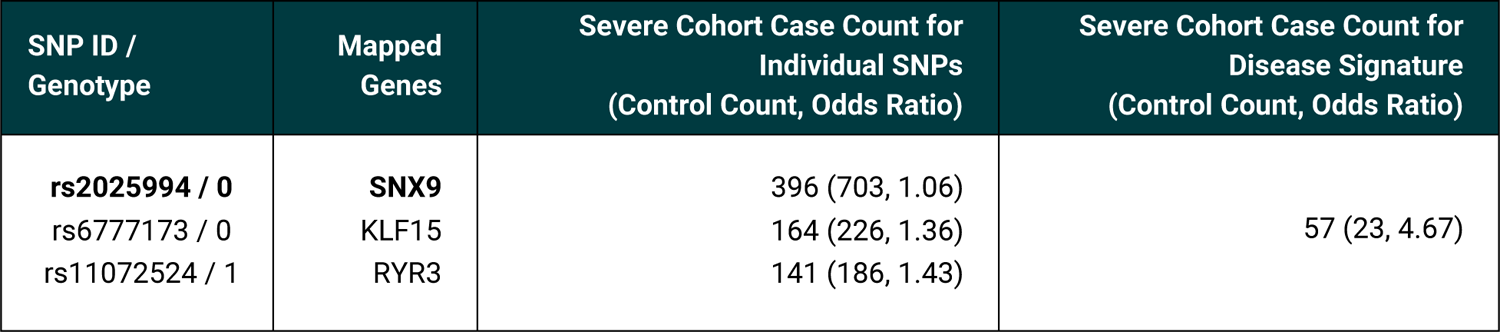
Example of one of the combinatorial disease signatures identified by the PrecisionLife combinatorial analysis of the Severe long COVID cohort. **Bold** text indicates the critical (RF-scored) SNPs (and the genes to which they are mapped) in this signature.

All cases included in the analysis possessed at least one of the disease signatures found to be significant in the hypothesis-free study of its cohort. The complete list of genetic variants and their mapped genes identified from this study are listed in Supplementary File 2.

Upon further evaluation, 118 (10%) disease signatures identified in the Severe cohort and 120 (8.4%) signatures in the Fatigue Dominant cohort comprised of SNPs that could be mapped to genes with shared biological functions or pathways (see Supplementary File 3).

As there were limited number of cases and controls of non-European ancestry (see Supplementary Table 6) in each of the two datasets, we evaluated the output to identify any disease signatures that may be confounded by population structure effects rather than reflecting a true disease signal.

All disease signatures in the Severe cohort passed the ancestry confounder analysis. We identified 129 (9%) disease signatures in the Fatigue Dominant cohorts that did not pass the ancestry confounder check (Supplementary Table 7). However, when we removed the SNPs and mapped genes represented only by these potentially confounding disease signatures (and not also by one or more additional true disease signatures), only one gene (*AC005005.1*) associated with the Fatigue Dominant cohort linked to the critical SNP, rs4820946, was eliminated from all final disease associated gene lists. This reduced the 74 genes found to 73.

The cohort analysis indicates that fewer than 15% of cases that were assigned to either one or both long COVID case groups, were hospitalized with severe COVID-19 or reported co-associated chronic diseases such as diabetes, cardiovascular disease or cognitive impairment. This meant that the number of cases with these phenotypes was too low to identify any associations, such as COVID-19 severity or a particular comorbidity, with genetic disease signatures.

Enrichment analysis of the fatigue, respiratory and mental health symptom-based scores for the Severe long COVID patients was used to investigate the clinical characteristics of the disease signatures identified in the Severe cohort study. Unfortunately, the population sizes were too small to reach statistical significance (*p* <0.05) after multiple-testing correction (see Supplementary File 4).

From the two independent hypothesis-free analyses of the datasets, we identified SNP genotypes mapping to 5 genes that were found to be significantly associated with disease in both the Severe and Fatigue Dominant long COVID cohorts. For each gene, more than 70% of cases from both cohorts possessed at least one disease signature containing an associated SNP (Table 4). These genes have a range of different functions and potential mechanism of action hypotheses as to their role in the development of long COVID.

**Table 4:** List of genes significantly associated with long COVID in both the Severe and Fatigue Dominant cohorts.

| Gene | % patients with corresponding disease signature in Severe cases (Severe controls) | % patients with corresponding disease signature in Fatigue cases (Fatigue controls) | Gene Function | Mechanism of Action Hypothesis in Long COVID |
| --- | --- | --- | --- | --- |
| **D2HGDH** | 90.6 (12.5) | 70.2 (1.5) | Catalyzes the oxidation of D-2-hydroxyglutarate (D-2-HG) to alpha-ketoglutarate | Involved in mitochondrial functioning, also exhibits anti-inflammatory effects<sup>28</sup> |
| **GUCY1A2** | 82.1 (7.9) | 71.9 (1.6) | Guanylate cyclase, catalyzes the conversion of GTP to 3',5'-cyclic GMP and pyrophosphate | Downregulated (hub gene) in SARS-CoV-2 infection<sup>29</sup> |
| **PCSK2** | 93.9 (32.9) | 94.9 (10.6) | Proprotein convertase subtilisin, processes hormones, involved in glucagon release | Maintains energy homeostasis, regulates circulating GLP-1 levels<sup>30</sup>. Blood glucose, insulin resistance and diabetes associated with long COVID<sup>31</sup> |
| **CCDC146** | 92.4 (15.2) | 86.8 (3.7) | Coiled-coil domain containing 146, a ubiquitous centriole and microtubule-associated protein | Associated with cognitive functioning and type 2 diabetes<sup>32</sup> |
| **PGPEP1** | 82.4 (34.0) | 91.0 (5.7) | Removes 5-oxoproline from various penultimate amino acid residues | Novel, possibly regulates various hormones and neuropeptides |

### Seeded Analysis to Test Overlap between Long Covid Cohorts

The two independent analyses of the Fatigue Dominant and Severe cohorts indicated that 5 genes were strongly associated with long COVID in both cohorts. We performed two seeded analyses to understand if any additional genes identified in either the Fatigue or Severe cohorts were also significant in the other population.

This approach revealed that 28 / 43 genes identified in the Severe cohort were also significantly associated with disease in the Fatigue Dominant cohort, and 25 / 35 genes from the original Fatigue Dominant analysis were also associated in the Severe cohort. This left 15 genes unique to the Severe cohort and 10 genes unique to the Fatigue Dominant cohort. The unique genes, the percentage of total cases they were associated with, and their biological functions are summarized in Tables 5 and 6.

**Table 5:** List of genes that were uniquely associated with the Severe case cohort.

| Gene | % patients with corresponding disease signature in Severe cases (Severe controls) | Gene Function | Mechanism of Action Hypothesis in Long COVID |
| --- | --- | --- | --- |
| **ADIPOQ** | 67.1 (20.6) | Adiponectin | Controls fat metabolism and insulin sensitivity<sup>33</sup><br>Prevents SARS-CoV2-induced acute lung injury<sup>34</sup> |
| **C1orf50** | 88.0 (9.6) | Chromosome open reading frame 50 | Novel |
| **CETP** | 72.3 (5.6) | Cholesteryl ester transfer protein | Role in insulin resistance, metabolic syndrome, macrophage-induced inflammation<sup>35</sup> |
| **CPLX4** | 71.2 (4.8) | Complexin 4 | Novel |
| **DLC1** | 26.8 (3.3) | GTPase, deleted in liver cancer 1 | Autophagy<sup>36</sup>, oncogene |
| **DSCAML1** | 47.3 (2.3) | Down syndrome cell adhesion molecule like 1 | Regulates corticotropin-releasing hormone in HPA axis, attenuated response to acute stressors<sup>37</sup> |
| **ENSG00000283580** | 88.02 (9.6) | Novel protein | Novel |
| **ENSG00000285082** | 52.3 (6.5) | Uncharacterized protein | Novel |
| **ETS1** | 30.5 (1) | Transcription factor, v-ets avian erythroblastosis virus E26 oncogene homolog 1 | Differentially regulated in peripheral blood of severe COVID-19 patients, modulates cytokine response<sup>38 39 40</sup> |
| **MARCH8** | 74.5 (5.8) | Membrane-associated ring finger, ubiquitin protein ligase | Downregulates host transmembrane protein, confers resistance to multiple viruses including SARS-CoV<sup>41 42</sup> |
| **NOL4** | 37.7 (7.6) | Nucleolar protein 4 | Differentially expressed in infective endocarditis<sup>43</sup> |
| **PDE6C** | 39.7 (8.3) | Phosphodiesterase 6C, cGMP-specific | Novel |
| **PGPEP1** | 82.4 (34) | Pyroglutamyl-peptidase I | Novel |
| **SNX9** | 41.2 (9.7) | Sorting nexin 9 | Regulated by chronic inflammation, trafficking of mitochondrial-derived vesicles<sup>44 45</sup> |
| **TLR4** | 52.3 (6.5) | Toll-like receptor 4 | Mediates innate immune response, genetic link to long-term cognitive dysfunction post COVID-19<sup>46 47</sup> |

**Table 6:** List of genes that were uniquely associated with the Fatigue Dominant case cohort.

| Gene | % patients with corresponding disease signature in Fatigue cases (Fatigue controls) | Gene Function | Mechanism of Action Hypothesis in Long COVID |
| --- | --- | --- | --- |
| **ABCA9** | 74.6 (1.5) | ATP-binding cassette | Cholesterol responsive gene involved in monocyte differentiation<sup>48</sup> |
| **ACOT12** | 14.3 (1.6) | Acyl-coA thioesterase | Acetyl-coA signaling and cholesterol biosynthesis<sup>49</sup> |
| **ANKRD6** | 25.6 (1.1) | Ankyrin repeat domain 6 | Possible links to muscle function and lipid metabolism<sup>50,51</sup> |
| **LYRM2** | 25.6 (1.1) | LYR motif containing 6 | Assembly of NADH-dehydrogenase complex, involved in cellular respiration<sup>52</sup> |
| **POR** | 23.9 (2.2) | Cytochrome P450 oxidoreductase | Downstream of MAPK signaling in oxidative stress pathway<sup>53</sup> |
| ***RRBP1*** | 71.2 (17.6) | Ribosome binding protein 1 | Relocates to mitochondrial vicinity during mitochondrial protein import stress, involved in endurance capacity in skeletal muscle during exercise<sup>54</sup> |
| ***SPTBN5*** | 72.3 (1.54) | Spectrin, beta, non-erythrocytic 5 | Novel |
| ***TNIK*** | 74.4 (19.1) | TRAF2 and NCK interacting kinase | Regulates JNK signaling<sup>55</sup> |
| ***TNS1*** | 83.2 (2.97) | Tensin 1 | Lack of AMPK increases tensin expression<sup>56</sup> |
| ***TPST1*** | 84.5 (2.64) | Tyrosylprotein sulfotransferase 1 | Required for monocyte recruitment<sup>57</sup> |

### Comparison of Long COVID with ME/CFS

We also used the seeded analysis approach to test for overlap between disease signatures associated with long COVID and those associated with ME/CFS in our previous study^16^.

Taking the list of SNPs within genes that were identified to be significant within the UK Biobank ME/CFS population, we found that 24 SNPs were also associated with long COVID in the Severe cohort. Of these 24 SNPs, 9 were critical (RF scored) within the Severe long COVID population, mapping to 5 genes (Table 7).

**Table 7:** List of critical SNPs significantly associated with long COVID in the Severe and Fatigue Dominant long COVID cohorts that can be linked to genes identified in a combinatorial analysis of UK Biobank ME/CFS patients.

| Genes identified in UK Biobank ME/CFS study | Critical SNP identified in Severe cohort (within 10 kb up/downstream of gene) | Critical SNP identified in Fatigue Dominant cohort (within 10 kb up/downstream of gene) |
| --- | --- | --- |
| **CLOCK** | - | rs62303689 |
| **SLC15A4** | rs11059915 | rs11059915 |
| **GPC5** | - | rs1536620 |
|  | - | rs16946160 |
|  | rs462954 | - |
|  | rs9560843 | - |
|  | rs989236 | - |
|  | rs9301839 | - |
| **ATP9A** | - | rs6096573 |
|  | rs77771672 | rs77771672 |
|  | rs2426361 | - |
| **INSR** | rs8110533 | rs8110533 |
| **USP6NL** | rs11257114 | - |

In the Fatigue Dominant cohort, 27 SNPs were associated with long COVID, of which 12 SNPs were also common with the Severe cohort (Supplementary Table 4). 7 of these 27 SNPs were critical (RF scored) SNPs within the Fatigue Dominant long COVID cases, mapping to 5 genes previously found in the ME/CFS study (Table 7).

### Comparison of Long COVID Genes Identified with Acute COVID-19 Studies

Whilst few GWAS significant variants have so far been identified in long COVID^58^, we sought to compare the 73 unique genes identified in our long COVID studies against the literature for any evidence within severe COVID-19 and/or long COVID. Of these genes, at least 9 have prior associations – such as differential expression and genetic susceptibility analyses - to acute COVID-19 after reviewing available publications in PubMed and other data sources such as OpenTargets (Table 8).

**Table 8:**
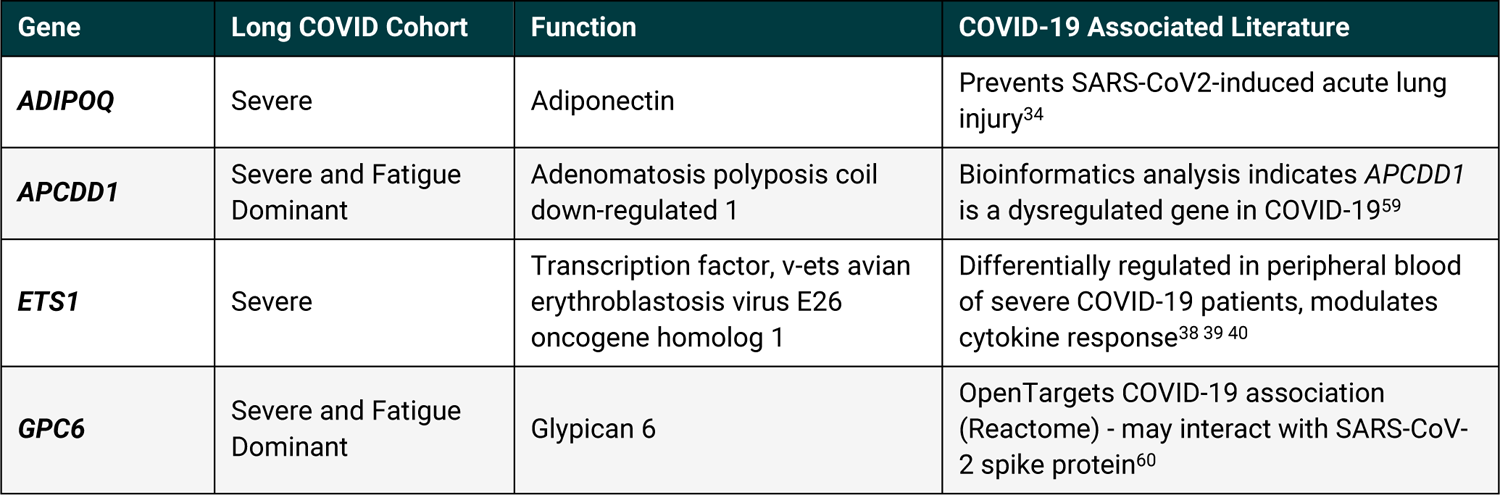
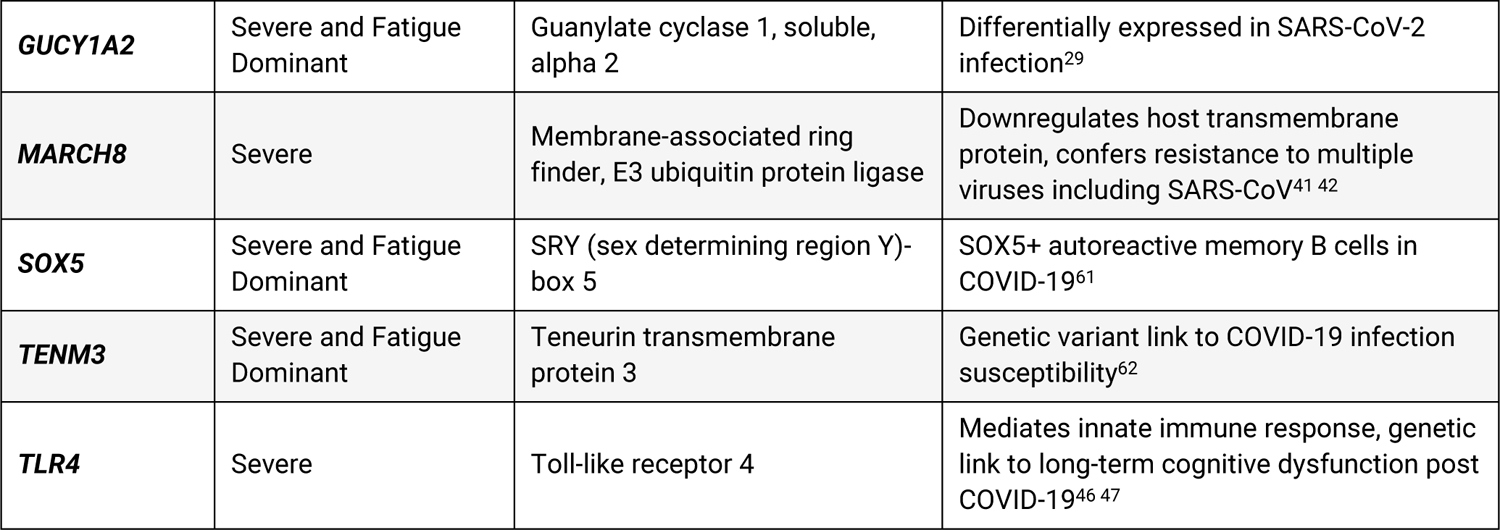
Expanded Genotypes Analysis results for 5 RF-scored genes identified in the Severe cohort linked to disease signatures of 2 or 3 SNPs (one disease signature contains SNPs associated with two genes).

| Gene | Long COVID Cohort | Function | COVID-19 Associated Literature |
| --- | --- | --- | --- |
| **ADIPOQ** | Severe | Adiponectin | Prevents SARS-CoV2-induced acute lung injury<sup>34</sup> |
| **APCDD1** | Severe and Fatigue Dominant | Adenomatous polyposis coil down-regulated 1 | Bioinformatics analysis indicates APCDD1 is a dysregulated gene in COVID-19<sup>59</sup> |
| **ETS1** | Severe | Transcription factor, v-ets avian erythroblastosis virus E26 oncogene homolog 1 | Differentially regulated in peripheral blood of severe COVID-19 patients, modulates cytokine response<sup>38 39 40</sup> |
| **GPC6** | Severe and Fatigue Dominant | Glypican 6 | OpenTargets COVID-19 association (Reactome) - may interact with SARS-CoV-2 spike protein<sup>60</sup> |
| **GUCY1A2** | Severe and Fatigue Dominant | Guanylate cyclase 1, soluble, alpha 2 | Differentially expressed in SARS-CoV-2 infection<sup>29</sup> |
| **MARCH8** | Severe | Membrane-associated ring finder, E3 ubiquitin protein ligase | Downregulates host transmembrane protein, confers resistance to multiple viruses including SARS-CoV<sup>41 42</sup> |
| **SOX5** | Severe and Fatigue Dominant | SRY (sex determining region Y)-box 5 | SOX5+ autoreactive memory B cells in COVID-19<sup>61</sup> |
| **TENM3** | Severe and Fatigue Dominant | Teneurin transmembrane protein 3 | Genetic variant link to COVID-19 infection susceptibility<sup>62</sup> |
| **TLR4** | Severe | Toll-like receptor 4 | Mediates innate immune response, genetic link to long-term cognitive dysfunction post COVID-19<sup>46 47</sup> |

**Table 8:** Known associations of genes identified in either one or both of the cohorts of long COVID patients with acute COVID-19.

| Target Gene | *SNX9* | *ADIPOQ* | *DLC1* | *NOL4* | *PDE6C* |
| --- | --- | --- | --- | --- | --- |
| Universally causative minor allele | 0 | 0 | 0 | 0 | 0 |
| Universally protective minor allele | 0 | 3 | 0 | 2 | 2 |
| Minor allele with SNP-specific causative effect | 2 | 0 | 0 | 0 | 0 |
| Minor allele with SNP-specific protective effect | 2 | 3 | 0 | 0 | 2 |
| Minor allele with combination-specific causative effect | 0 | 3 | 0 | 2 | 0 |
| Minor allele with combination-specific protective effect | 0 | 0 | 0 | 0 | 0 |
| Ambiguous / no consistent effect | 0 | 1 | 2 | 0 | 0 |

We also compared our results against the blood derived gene expression signatures associated with post-acute sequelae identified by Thompson *et al*^63^. There are several key differences between the studies – Thompson *et al* recruited individuals hospitalized with severe acute COVID-19 infection, whereas the majority of individuals in our study experienced milder forms of the disease (Table 1). We are also drawing comparisons from a transcriptomic study derived from whole blood against a combinatorial study of germline genetic variants.

Nonetheless, we found that 14 of the 73 genes (Severe=7 and Fatigue Dominant=7) identified in our analyses were also differentially expressed at the transcriptomic level in patients experiencing long COVID (Supplementary Table 10).

### Overlap Between Long COVID and Other Diseases

We identified genes with known genetic associations across a wide range of complex diseases including neurodegenerative, mental or behavioral, cardiovascular, gastrointestinal, autoimmune and metabolic diseases (see Supplementary Tables 8 and 9). We evaluated the degree of overlap at a biological process level (using mapping of genes to biological processes in Gene Ontology^64, 65^) to identify the common pathophysiological mechanisms that are shared between those diseases and long COVID.

27 biological processes are significantly enriched in the 73 long COVID genes identified in this analysis, of which 19 processes are also significantly enriched in at least one other indication group (Supplementary Table 13). Based on these 19 pathways, long COVID genes shared the greatest number of biological processes (>50%) with cardiovascular disease and mental or behavioral disease followed by gastrointestinal disease, neurodegenerative disease, autoimmune disease and metabolic disease (Figure 7, Supplementary Table 13).

**Figure 5:**
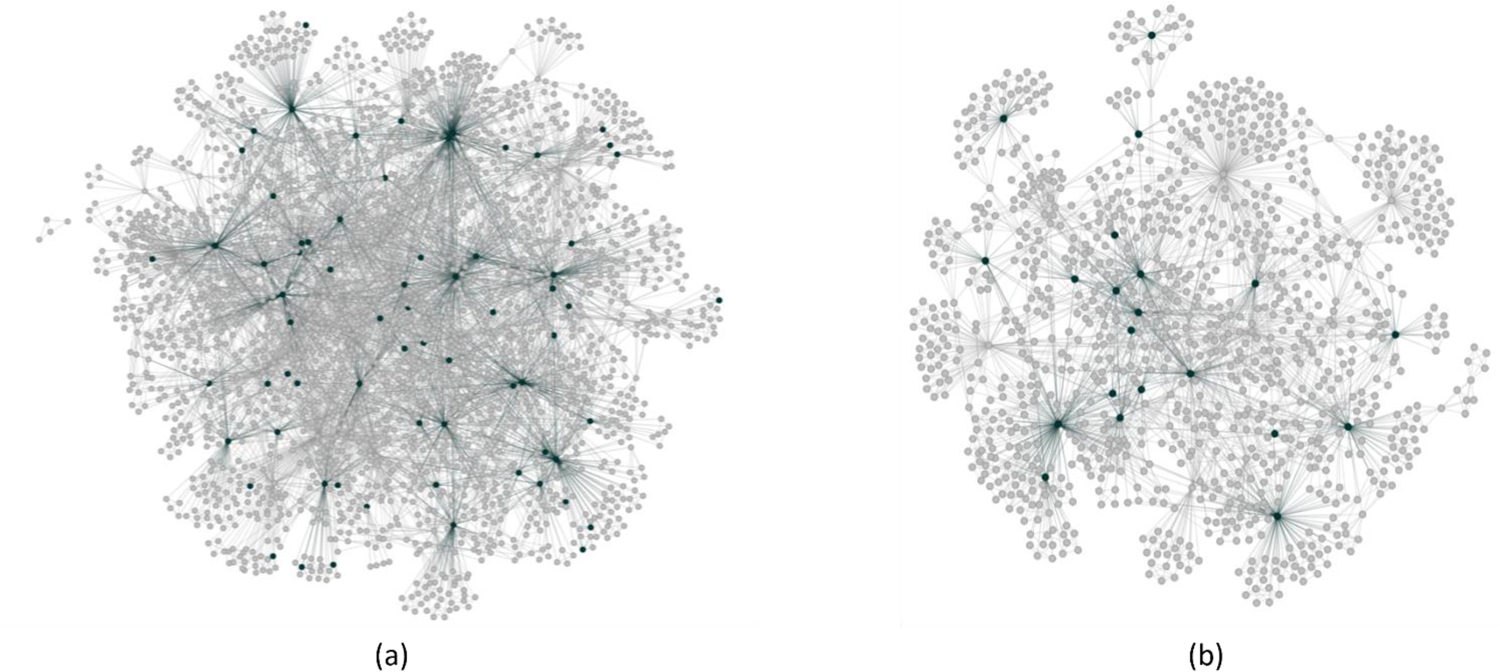
Disease architecture diagrams representing (a) the Severe and (b) Fatigue Dominant long COVID patient populations generated by the PrecisionLife platform. Each circle represents a disease-associated SNP genotype, and edges represent their co-association in patients in disease signature(s). The critical SNP genotypes identified in each case population are highlighted in dark green.

**Figure 6:**
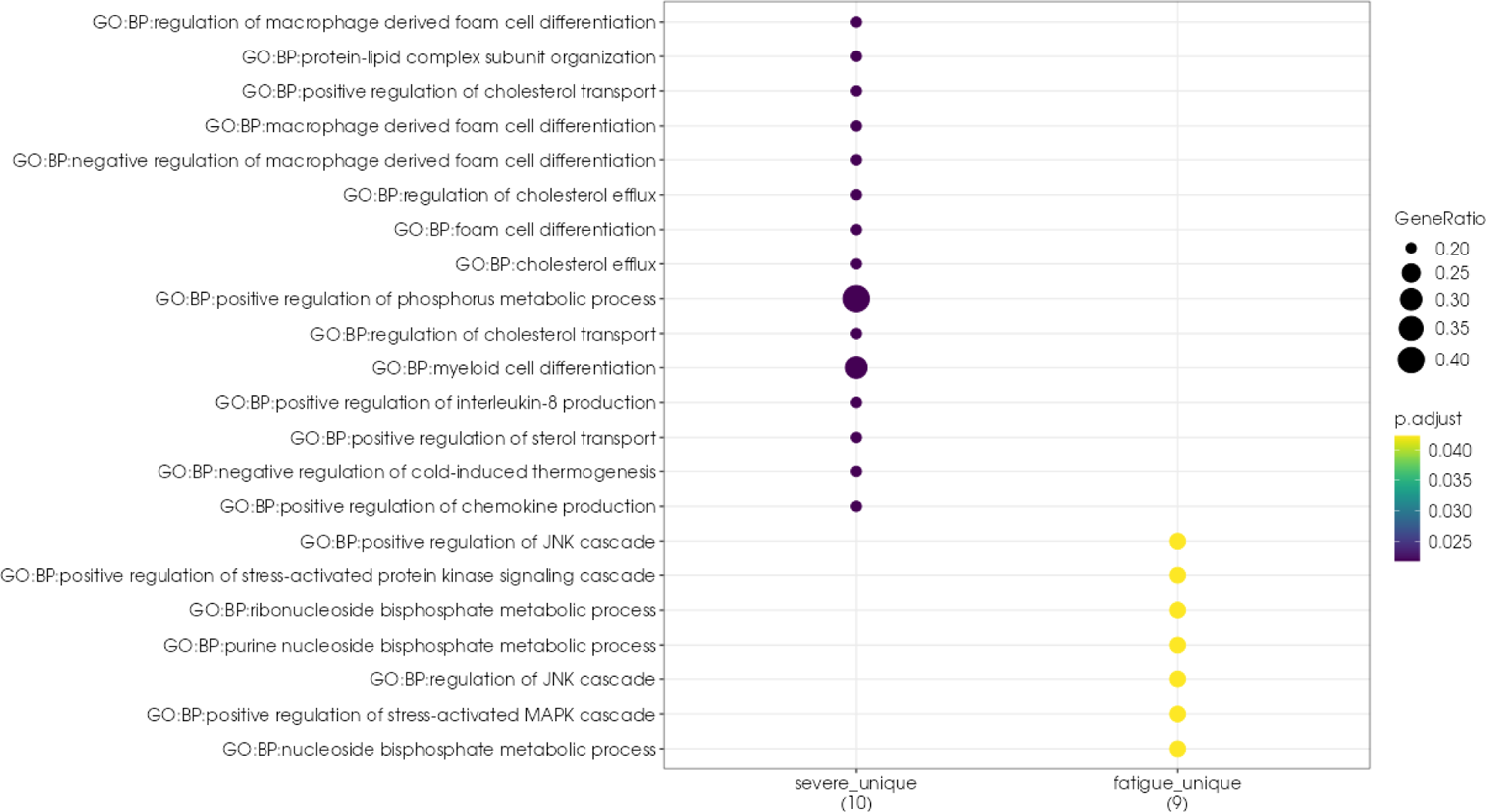
Pathway enrichment plot for disease-associated genes found in the Severe and Fatigue Dominant long COVID cohorts. GeneRatio represents the ratio of genes found in the pathway compared to the genes associated with a cohort and *p*.adjust represents the *p*-value adjusted for multiple testing. The dots in the plot are colour-coded based on their corresponding *p*.adjust values.

**Figure 7:**
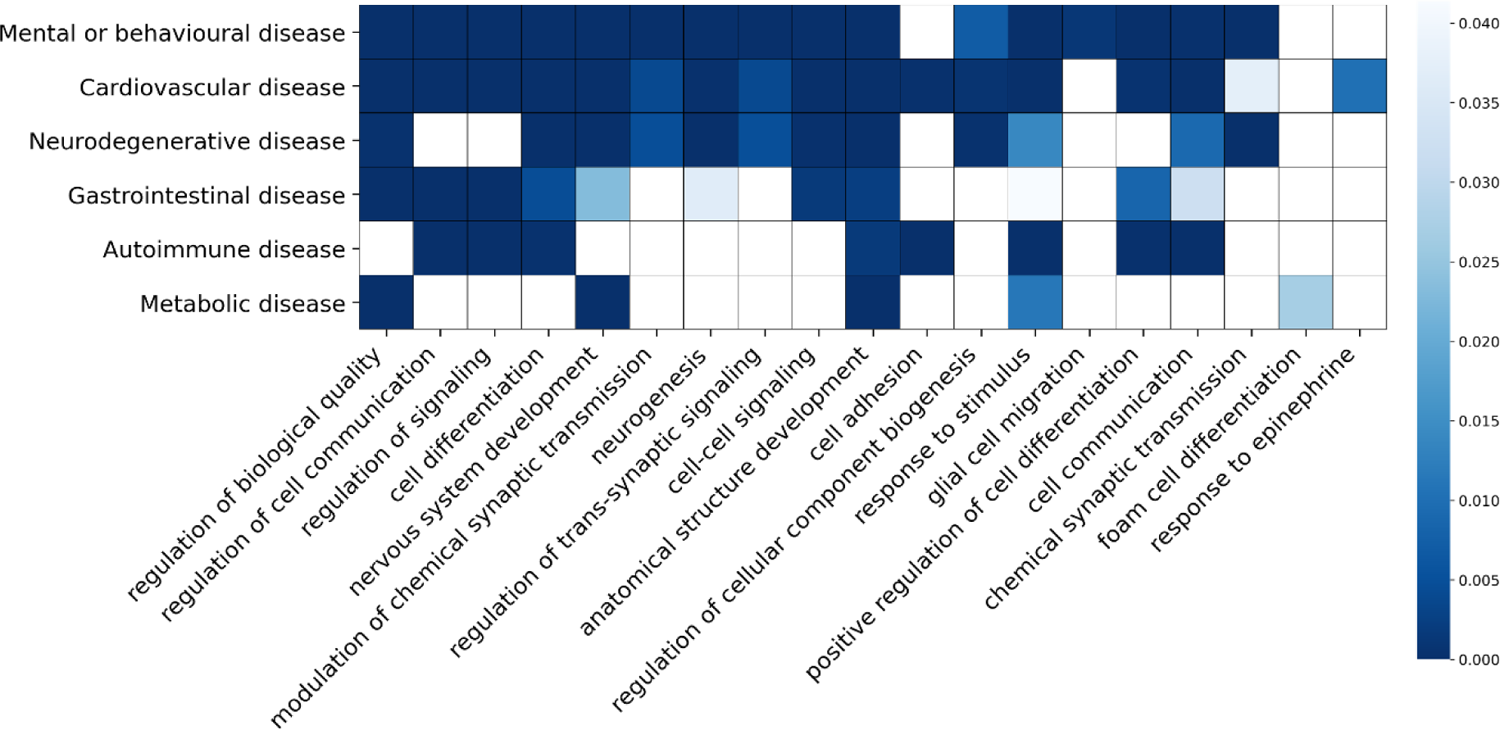
Heatmap plot showing 19 biological processes (Gene Ontology biological process terms) shared between 73 long COVID genes identified in the GOLD cohort and genes with genetic evidence in one or more indication groups (neurodegenerative, mental or behavioral, cardiovascular, gastrointestinal, autoimmune and metabolic disorders). For each indication group, only the significantly enriched biological processes (*p* < 0.05) are shown in blue and the intensity of the color is based on the *p* values of the Gene Ontology term in each indication group.

### Expanded Genotypes Analysis to detect Causal Features

We conducted expanded genotypes analysis for all Severe cohort RF scored genes (see Tables 3 and 4) found in disease signatures with 2 or 3 SNP genotypes. These comprise 5 genes corresponding to 23 disease signatures, including a disease signature that contains two RF scored genes (see Table 8).

We found that the critical SNP is universally protective across at least 2 validated disease signatures for 3 of the 5 RF scored genes (*ADIPOQ, NOL4, and PDE6C*). That is, when we control for the genotypes at the interacting SNPs, expanded genotype signatures featuring at least one copy of the critical SNP minor allele are consistently associated with lower odds of severe long COVID relative to expanded genotype signatures with the homozygous wild type genotype for the critical SNP. In all but one of the remaining disease signatures for these genes, the critical SNP minor allele is most often associated with decreased odds of severe long COVID, with narrow exceptions: i.e., when it fails to co-occur with the minor allele of an interacting SNP (“SNP-specific protective effect”) or when it co-occurs with a specific set of genotypes at multiple interacting SNPs (“combination-specific causative effect”).

The critical SNP minor alleles for these three genes are typically associated with decreased risk of severe long COVID, which either implies that they represent broadly protective variants or causative variants that are in LD with the wild type allele at the genotyped SNP. This relationship only becomes apparent, however, when we control for the confounding effects of other causative and/or protective variants. Only one validated disease signature for these three genes fails to exhibit a consistent biological association between the critical SNP minor allele and disease, indicating a potential false positive.

In contrast, the gene *SNX9* consistently is associated with more complex interactions that highlight the combinatorial dynamics of disease. For example, we identified a disease signature comprising three SNPs that is associated with strongly elevated odds of long COVID. This disease signature includes:

- critical SNP rs2025994 located approximately 40kb upstream of the *SNX9* coding region
- interacting SNP rs6777173 located 12 kb upstream of *KLF15*
- interacting SNP rs11072524 located in an intron of *RYR3*

We found that the *SNX9* minor allele offers significant protection against the risk of long COVID among patients who possess a copy of the minor allele at either interacting SNP (i.e., a SNP-specific protective effect). That is, patients with the *SNX9* heterozygous or homozygous minor allele genotype consistently have lower odds of developing severe long COVID than patients with the *SNX9* homozygous wild type genotype, after controlling for the confounding effects of the genotypes at the two interacting SNPs (see Table 9). Due to the small sample sizes associated with many expanded genotype signatures, these individual comparisons are not statistically significant. However, if we pool all patients in this cohort, then patients with a copy of the *SNX9* minor allele have significantly lower odds of disease than patients who are homozygous for the *SNX9* wild type allele (odds ratio = 0.52, 41 cases / 134 controls vs. 316 cases / 532 controls, Fisher’s Exact Test *p*=0.00047; note that these totals include patients with rare expanded genotype signatures not shown in Table 9).

**Table 9:** Assessing the effects of the *SNX9* rs2025994 genotype on severe long COVID when controlling for the genotypes of the interacting SNPs rs6777173 (*KLF15*) and rs11072524 (*RYR3*). We present comparisons for genotype combinations that are present in more than 10 patients (which excludes the 18 patients who are homozygous for the *SNX9* minor allele). None of the EGA odds ratios for the individual comparisons are statistically significant after correcting for multiple testing. However, among patients who possess a copy of the minor allele at either interacting SNP, presence of the *SNX9* minor allele consistently results in lower odds of disease relative to the homozygous wild type genotype (Fisher’s Exact Test *p*=0.00047). Among patients who possess only wild type alleles for both interacting SNPs, presence of the *SNX9* minor allele results in higher odds of disease relative to the homozygous wild type genotype, but the difference is not statistically significant (Fisher’s Exact Test *p*=0.075).

| *KLF15*<br>minor allele<br>count | *RYR3*<br>minor allele<br>count | *SNX9* homozygous wild type<br>Odds<br>(Cases:Controls) | *SNX9* heterozygous<br>Odds<br>(Cases:Controls) | EGA Odds Ratio: *SNX9*<br>heterozygous vs.<br>homozygous wild type<br>(95% confidence<br>interval) |
| --- | --- | --- | --- | --- |
| 0 | 0 | **0.46**<br>(74:160) | **0.89**<br>(17:19) | **1.93**<br>(0.95 – 3.93) |
| 0 | 1 | **2.48**<br>(57:23) | **0.70**<br>(7:10) | **0.28**<br>(0.10 – 0.83) |
| 1 | 0 | **0.43**<br>(114:267) | **0.28**<br>(14:50) | **0.66**<br>(0.35 – 1.23) |
| 1 | 1 | **0.63**<br>(48:76) | **0.24**<br>(6:25) | **0.38**<br>(0.15 – 0.99) |
| 2 | 0 | **0.66**<br>(71:108) | **0.21**<br>(6:28) | **0.33**<br>(0.13 – 0.83) |
| 2 | 1 | **0.49**<br>(17:35) | **0.38**<br>(3:8) | **0.77**<br>(0.18 – 3.29) |

A different pattern arises among patients who are homozygous for the wild type genotype at both interacting SNPs. Here, patients with a copy of the *SNX9* minor allele have higher odds of disease than patients who are homozygous for the *SNX9* wild type allele (odds ratio = 1.86, 19 cases / 22 controls vs. 74 cases / 160 controls), although the odds ratio is not statistically significant (Fisher’s Exact Test *p*=0.075).

Together these results suggest that the *SNX9* genotype is a significant contributor to the risk of severe long COVID infection, but that the gene-disease relationship is context dependent and mediated by interactions with *KLF15* and *RYR3*. Similar non-linear interactions are represented by three additional disease signatures comprised of the same *SNX9* critical SNP and different interacting SNPs. Monogenic approaches such as GWAS that do not consider these gene-gene interactions can fail to detect potentially important drivers of disease.

Finally, the expanded genotypes analysis did not provide any additional insight into the relationship between *DLC1* and disease. This could indicate that the biological relationship between *DLC1* is highly complex or that the result is a false positive. However, the disease signatures associated with strongly elevated odds of severe long COVID all contain the rare homozygous minor allele genotype for the *DLC1* critical SNP. Due to small sample sizes, we were unable to analyze other expanded genotype signatures containing the potentially causative genotype. Thus, the ambiguous results may reflect the fact that the relationship between the *DLC1* minor allele and long COVID does not carry over into heterozygous patients.

### Evaluation of Potential Novel Drug Targets and Repurposing Opportunities

We evaluated the genes identified in the study to find potential novel drug targets and their associated mechanistic patient stratification biomarkers (the disease signatures that connect patient subgroups with the mechanistic etiology for their disease). As described in our previous ME/CFS paper, the use of combinatorial analytics to identify novel targets has been validated in other diseases such as ALS, where these novel targets have demonstrated disease modifying activity in *in vitro* models^66^.

Of the 73 unique genes found across the two cohorts, 42 are potentially tractable targets for drug development strategies based on annotations from OpenTargets (defined by a score of greater than 0 across at least one metric for tractability), see Supplementary Table 11. This includes 26 targets that are suited to an antibody approach and 18 that are amenable to modulation by small molecules.

Most (> 90%) of the genes are expressed in a wide range of tissues (Supplementary Figure 6) although the expression profile of the genes in specific cell types is variable (Supplementary Figure 7). Approximately 44% (*n*=30) of the genes are expressed in inhibitory neurons followed by 41% in excitatory neurons (*n*=28) and 40% in oligodendrocyte precursor cells (n=27).

Using a systematic repositioning approach^67^, we identified 13 long COVID targets that already have drugs in clinical development. As these drugs or development candidates may require fewer preclinical studies and already have a known safety profile, they could represent a quicker and de-risked strategy for developing potential new treatments. We are exploring the repurposing potential of these compounds for the treatment of long COVID and ME/CFS (where appropriate).

From this analysis for example, we identified TLR4 as an attractive repurposing candidate. Our analysis indicates that 52% of cases included the Severe long COVID cohort had at least one disease signature containing a variant containing TLR4 and there is additional supporting evidence that inhibition of TLR4 in a mouse model prevents long term cognitive pathology such as synapse elimination and memory deficits that is caused by the SARS-CoV-2 Spike protein^46^. Clinical studies have already shown that antagonizing TLR4 signaling dampens the pathological cytokine storm observed in patients with severe acute COVID-19 and reduces mortality rates in hospitalized COVID-19 patients^68, 69^. However, our analysis also indicates that antagonism of TLR4 may demonstrate therapeutic effects in long term pathology caused by SARS-CoV-2.

We performed a search of the GlobalData^70^ database to further understand the number and stage of development of TLR4 antagonists that are in clinical pipelines. This revealed a total of 88 unique drugs that target TLR4 (either singularly or as part of a combination therapy), including 8 in development for acute COVID-19, the most advanced of which (Paridiprubart, Edesa Biotech Inc) is currently being evaluated in a Phase 3 study in hospitalized COVID-19 patients with Acute Respiratory Distress Syndrome (ARDS)^71^.

## Discussion

As an approach to identify the drivers of the complex disease biology of long COVID, combinatorial analytics yields more useful signal than GWAS. No SNPs reached the genome-wide significance threshold in either the Severe or Fatigue Dominant cohorts. This underlines the difficulties involved in using monogenic analysis approaches to understanding disease associated genetic variants and mechanistic etiologies in heterogeneous and polygenic diseases, especially with small datasets.

Using combinatorial analytics, we identified 73 unique genes in a long COVID population and highlighted the relevance of subsets of these genes to the different sub-cohorts of the disease population. At least 9 of the genes identified in this study have been linked to acute COVID-19, and despite key differences in the study designs, we also observe that 14 of the 73 genes were differentially expressed in a transcriptomic analysis of long COVID patients. We can form strong mechanism of action hypotheses for each gene’s role in the development of long COVID.

Splitting the population into two long COVID subtypes, Severe and Fatigue Dominant, allowed us to explore the genetic and biological differences underpinning different clinical manifestations. The comparative pathway enrichment analysis identified differences in pathways between the genes uniquely associated with the Severe long COVID group and those uniquely associated with the Fatigue Dominant phenotype (Figure). The greater number of genes involved in immune response in the Severe long COVID cohort may also indicate a more severe form of the acute infection. This may potentially arise as a result of patients experiencing higher viral loads than average, as we identified 4 genes that have been functionally linked to SARS-CoV-2 host response and/or acute severe COVID-19 (Table 5).

The pathway enrichment analysis also highlighted an overrepresentation of genes involved in macrophage foam cell differentiation. The formation of foam cells leading to a profibrotic macrophage phenotype is critical in the development of atherosclerosis^72^. However, there is also evidence that profibrotic pulmonary macrophages contribute to acute respiratory distress syndrome (ARDS) and lung injury associated with patients with severe COVID-19^73^.

The genes that were associated only with the Fatigue Dominant long COVID cohort are enriched in MAPK and JNK signaling cascades as well as other metabolic processes involved in mitochondrial function and cellular respiration (Table 6). As discussed in our previous ME/CFS paper, dysregulated mitochondrial function, resulting in the inability to increase respiration rates in response to increased demand from stressors such as exercise^74^, may result in the post-exertional malaise (PEM) that is a hallmark of ME/CFS. The finding of similar pathways in the Fatigue Dominant long COVID cohort suggests that these patients may also struggle to meet energy demands.

It is known that NK cell effector function (cytotoxic activity) regulated by MAPK signaling cascades, including via the c-Jun N-terminal kinase (JNK)^75^ signaling pathway, is dysregulated within patients with ME/CFS, who exhibit reduced NK cell cytotoxic activity^76^. Further work will be required to confirm if similar pathological events occur in patients who develop fatigue dominant long COVID.

When we evaluated the degree of similarity between the genes associated with ME/CFS and long COVID, we found 13 critical SNPs (39 in total) within at least one of the long COVID populations that could be mapped to a gene previously associated with ME/CFS.

In both Severe and Fatigue Dominant long COVID populations, we identified SNPs mapping to the genes *ATP9A, INSR, CLOCK, SLC15A4* and *GPC5.* All of these genetic variants were found in a higher proportion of the Fatigue Dominant and Severe long COVID populations than in the ME/CFS case group. This finding may indicate that the long COVID case group defined by fatigue symptoms is more homogenous than those within the self-reported ME/CFS population, which likely includes a mix of viral and non-viral triggers of chronic fatigue symptoms.

We found that the *CLOCK* gene is significantly associated with Fatigue Dominant long COVID and ME/CFS. CLOCK (Circadian Locomotor Output Cycles Kaput) is an important regulator of circadian rhythm, disruptions of which have been associated with pain, insomnia, insulin resistance, immunological function and impaired mitochondrial function^77–81^. Interestingly, one of the most common variants identified in ∼86% of the long COVID Fatigue Dominant population mapped to the gene *NLGN1*. *NLGN1* is also transcriptionally activated by CLOCK in the forebrain^82^, which could indicate multiple genetic contributions to dysregulated circadian rhythm in long COVID.

Of the remaining 4 genes common between long COVID and ME/CFS, we identified 3 common variants in the genes *ATP9A, INSR* and *SLC15A4* in both Severe and Fatigue Dominant cohorts (Table 7).

*SLC15A4* encodes a transmembrane transport that has previously been associated with inflammatory autoimmune diseases such as systemic lupus erythematosus from genome-wide association studies^83, 84^. However, SLC15A4 also plays a key role in mitochondrial function, with knock down of the gene resulting in impaired autophagy and mitochondrial membrane potential under cell stress^85^.

We also hypothesized that the genetic variants in *ATP9A* and *INSR* both contribute to dysregulated insulin signaling in subgroups of ME/CFS patients. Type 2 diabetes-related signaling pathways and insulin resistance were also a key theme within the genes associated with long COVID, and 11 of the gene targets identified in this analysis have prior associations with type 2 diabetes in the OpenTargets database (Supplementary Table 12). Metabolic dysfunction and type 2 diabetes may increase risk of developing severe acute COVID-19^86^ and epidemiological studies have demonstrated that there is an increased risk of developing diabetes post COVID-19 compared against controls who had not been infected with SARS-CoV-2^87^. Furthermore, increased incidence of insulin resistance and glycemic dysregulation was observed in patients 2 months post COVID-19 and in long COVID patients^31, 88^.

Several of the biological processes that genes identified in this study are significantly enriched for – such as foam cell differentiation – are also associated with known genetic links to metabolic diseases such as type 2 diabetes (Figure). Metabolic dysfunction has a variety of biological consequences, including increased levels of chronic inflammation, dysregulated immune response to acute infection, endothelial cell dysfunction and defects in coagulation pathways. All of these have been linked to long COVID and severe acute COVID-19 pathogenesis^89^.

It is therefore plausible that patients with genetic variants that predispose them to metabolic dysfunction and insulin resistance are more likely to suffer from long term pathological sequelae after the acute phase of COVID-19 infection. From these findings we would indeed expect this population to have increased rates of new-onset type 2 diabetes compared to the non-long COVID population. Unfortunately, longitudinal health record data after the survey was completed was not available to validate this hypothesis in this analysis.

Similarities in indications observed from the cross-disease analysis have also highlighted shared pathways and biological processes associated with genetic drivers of these indications. The results are supported by common clinical manifestations reported in long COVID studies. Of the 27 pathways significantly enriched in the long COVID genes identified in this analysis, 16 (60%) are associated with gene targets previously associated with mental or behavioral disease (Figure). This includes indications such as major depressive disorder, anxiety disorder and schizophrenia. A recent meta-analysis of over 10,000 patients indicated that neurological and neuropsychiatric symptoms, such as brain fog, attention deficits and fatigue, were some of the most reported 3 months after acute COVID-19^90^. This analysis may indicate some of the genetic underpinnings of these manifestations post-COVID.

### Study Limitations

There are several limitations to this study. The most obvious is that the available datasets, even in a disease as topical, prevalent, and debilitating as long COVID are still very small, which notwithstanding the improved sensitivity offered by the combinatorial analytics approach, inevitably poses limits on the statistical power of the study.

The most challenging limitation is the poor representation of diverse ancestries, which is essential to gain a deeper understanding of the variability of disease etiology and achieve a level of health equity. As demonstrated by the cohort analysis, even though considerable effort was made to recruit as diverse a population as possible, the majority of participants recruited to the GOLD study were of self-reported white Caucasian ancestry. It is evident that long COVID is a highly heterogeneous disease with a variety of different symptoms, clinical presentations and underlying disease mechanisms including neurological and metabolic dysregulation. From this dataset, we cannot understand the varying prevalence of these symptoms, or the effects that different genetic ancestries, socioeconomic factors, pathogen exposure levels or geographical differences may have in influencing the risk and presentation of long COVID in different ancestries.

Our cohort analysis also revealed that the incidence of other comorbidities (such as type 2 diabetes, cardiovascular disease etc.) was lower than expected for a cohort with the same average age as the long COVID population. This may indicate a degree of ‘otherwise healthy’ volunteer bias that limits this dataset as a representative sample of long COVID. Alternatively, it could reflect a problem with under-reporting of other medical conditions within the self-reported questionnaire.

All the non-genomic data was self-reported by the participants via a questionnaire upon recruitment to the study, including long COVID symptoms, level of acute COVID-19 severity and medical history. Unfortunately, no further EHR/primary care data was available. This method for reporting the degree of long COVID symptoms experienced is likely to be more subjective and prone to memory lapses and retrospective interpretation than direct and concurrent clinical information. This creates challenges in identifying the most relevant clusters of long COVID symptoms (e.g., respiratory, fatigue, GI etc.) and evaluating the severity of those symptoms experienced by different subgroups of cases.

We were unable to fully evaluate some of the most significant consequences and secondary diagnoses associated with long COVID disease. In particular, we would have liked to evaluate the specific drivers underlying the development of POTS, which was only recorded as part of participants’ free-text responses and not captured in the main questionnaire. In the absence of consistent diagnosis and clinical reporting for POTS, we attempted to analyze the symptoms that patients reported when recruited to the study. Tachycardia, dizziness, palpitations, brain fog and even in some cases POTS were recorded but in insufficient numbers for a meaningful analysis.

Hospital admission with a more severe form of acute COVID-19 has previously been identified as a risk factor for the development of long COVID^91^. We were unable to test this finding, as fewer than 10% of any of our case cohorts were hospitalized with COVID-19. As a result, there was insufficient data available to explore if long COVID cases with the 9 variants mapped to genes previously associated with acute COVID-19 (Table 8) were more likely to have experienced a more severe form of acute COVID-19.

Finally, there is some emerging evidence that vaccination against COVID-19 may be protective against the development of long COVID^92^. The majority of cases included in our study were recruited in 2021 and the questionnaire did not contain any questions regarding vaccination status, or if the participants contracted acute COVID-19 before or after vaccination. As such, we are unable to evaluate the effect of vaccination on long COVID development within this cohort. There is also evidence that omicron variants are less likely to cause long-term symptoms even after adjusting for vaccine status^93^. However, it was also not possible to assess the association of SARS-CoV-2 variant status with long COVID risk.

## Conclusions and Future Perspectives

The results of this study, while encouraging and building consistently on findings in ME/CFS and other diseases with related symptomology, still need to be validated and replicated within an independent long COVID population, which ideally would have much deeper clinical phenotype and longitudinal history information.

Various groups have been collecting large acute COVID-19 and long COVID patient datasets over the last 3 years and we hope that they will now make the individual patient level data available to the wider research community quickly. We can realistically expect that analyzing an independent, larger and more detailed patient dataset using combinatorial analytics approaches will further improve the disease insights that we are gaining in long COVID, offering routes forward to alleviate the massive unmet medical need which has blighted the lives of millions of patients.

## Data Availability

All data produced in the present work are contained in the manuscript

## Acknowledgements

Research described in this article has been conducted using data from Sano Genetics’ Long COVID GOLD study and we thank the Sano team for their help in preparing these data. Special thanks to Anastasia Lankina and Mark Strivens who provided input into the manuscript, Gert Møller and Claus Erik Jensen, who initially developed the combinatorial analytics methodology, and the rest of the PrecisionLife team.

## Ethics Declarations

The Sano Genetics GOLD study has approval from the Wales Research Ethics Committee (REC) (IRAS 291221).

