## Supplementary Materials for "Genetic Risk Factors for Severe and Fatigue Dominant Long COVID and Commonalities with ME/CFS Identified by Combinatorial Analysis"

### Supplementary Data
#### Long COVID Manuscript

**Author(s):** Krystyna Taylor, Matthew Pearson, Sayoni Das, Jason Sardell, Karolina Chocian, Steve Gardner

**Date:** 13 July 2023

**Status:** v10

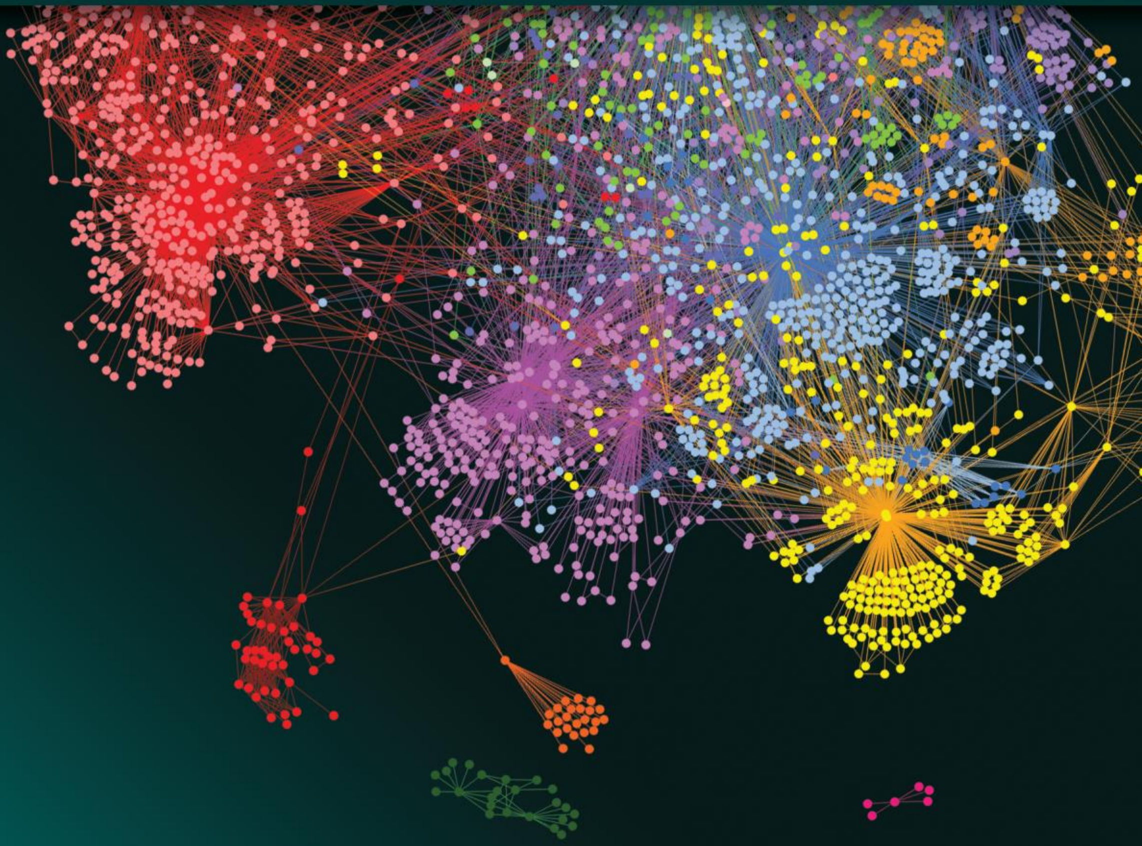

precisionlife.com

**PrecisionLife Ltd**
Registered Office: Unit 8b Bankside, Hanborough Business Park, Long Hanborough, OX29 8LJ
Company No. 08687703 VAT Number GB 172 5125 25

### 1 Supplementary File List

1. **Supplementary File 1**: Long COVID GOLD Study - Data Dictionary.xlsx

   The data dictionary for the Long COVID GOLD Study is available in this supplementary file.

2. **Supplementary File 2**: Long_COVID_studies_critical_snps.xlsx

   List of disease-associated critical SNPs and genes identified by the PrecisionLife platform in the three long COVID cohorts. Additional columns include gene associations from publicly available cell/tissue-specific eQTL and chromatin-interaction (Hi-C) data.

3. **Supplementary File 3**: Pathway enrichment of long COVID Disease Signatures.xlsx

   Pathway enrichment analysis results for disease signatures identified in Severe and Fatigue Dominant long COVID cohorts. The enrichment analysis was performed using g:Profiler using Gene Ontology, KEGG, Reactome and WikiPathways as data sources and considering only genes with at least one annotation as background genes. Only significant results ($p < 0.05$) after applying multiple testing correction were reported from the enrichment analysis.

4. **Supplementary File 4**: Severe Cohort Disease signatures associated with symptom-based scores.xlsx

   List of disease signatures that are correlated with symptom-based scores such as increase in respiratory, fatigue or mental health change scores prior to and after COVID.

### 2 Supplementary Information

#### 2.1 Sano Genetics Long COVID GOLD Study

**Supplementary Table 1:** Scores generated for respiratory, fatigue and mental health symptoms before and after COVID using participant questionnaire responses in the GOLD study.

| Symptom-based score | Participant reported scores derived from questionnaire responses | Attributes from questionnaire response used to calculate change in symptoms before and after COVID |
|---|---|---|
| **Respiratory Change** | Change in breathlessness at rest | breathlessness_rest_current - breathlessness_rest_pre_covid |
| | Change in breathlessness after dressing | breathlessness_dressing_current - breathlessness_dressing_pre_covid |
| | Change in breathlessness after climbing stars | breathlessness_climbing_stairs_current - breathlessness_climbing_stairs_pre_covid |
| **Fatigue Change** | Change in mobility problems | mobility_problems_severity_now - mobility_problems_severity_pre_covid |
| | Change in fatigue | fatigue_now - fatigue_pre_covid |
| | Change in personal care difficulties | personal_care_difficulties_now - personal_care_difficulties_pre_covid |
| | Change in difficulties to carry on usual activities | usual_activities_difficulty_now - usual_activities_difficulty_pre_covid |
| | Change in muscle pain or discomfort | muscle_pain_now - muscle_pain_pre_covid |
| **Mental Health Change** | Change in anxiety | anxiety_now - anxiety_pre_covid |
| | Change in depression | depression_now - depression_pre_covid |
| **Total Change** | Total change in Respiratory, Fatigue or Mental Health scores | Respiratory score + Fatigue score + Mental Health score |

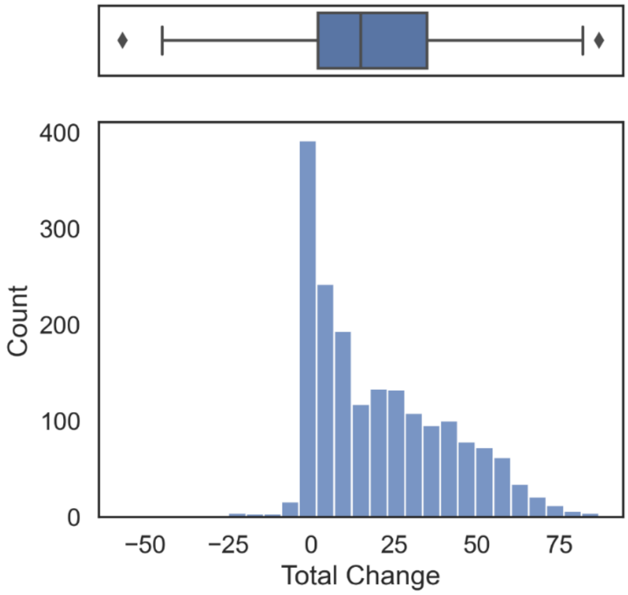

**Supplementary Figure 1:** Distribution of the 'Total Change' score for all individuals in the GOLD cohort.

**Supplementary Table 2:** List of top ranking (n=35) words in the free-text responses of 1,489 (81.41%) individuals in the GOLD study with a COVID diagnosis (n=1,829) along with Severe and Fatigue Dominant cases to the question on other symptoms individuals in the GOLD long COVID cohort have experienced since their illness that was not covered in the questionnaire. The rank of the words reflects their weights based on their incidence within each group.

| Rank | All GOLD study individuals with COVID diagnosis (*n*=1,828) | | Severe cases (*n*=459) | | Fatigue Dominant cases (*n*=477) | |
|---|---|---|---|---|---|---|
| **1** | **Word** | **Count** | **Word** | **Count** | **Word** | **Count** |
| **2** | smell | 125 | headache | 52 | pain | 62 |
| **3** | headache | 112 | tinnitus | 48 | headache | 57 |
| **4** | pain | 98 | pain | 46 | tinnitus | 53 |
| **5** | tinnitus | 97 | issue | 35 | issues | 37 |
| **6** | taste | 96 | POTS | 30 | symptom | 34 |
| **7** | issue | 74 | smell | 30 | POTS | 33 |
| **8** | problem | 70 | taste | 29 | smell | 33 |
| **9** | symptom | 65 | dizziness | 29 | dizziness | 31 |
| **10** | loss | 60 | symptom | 29 | problem | 31 |
| **11** | dizziness | 51 | problem | 28 | insomnia | 30 |
| **12** | insomnia | 51 | tachycardia | 27 | tachycardia | 30 |
| **13** | POTS | 43 | skin | 26 | eye | 28 |
| **14** | skin | 42 | loss | 26 | skin | 28 |
| **15** | palpitations | 41 | insomnia | 25 | taste | 27 |
| **16** | fatigue | 39 | eye | 24 | loss | 25 |
| **17** | hand | 39 | hand | 18 | hand | 22 |
| **18** | feeling | 36 | palpitations | 17 | palpitations | 19 |
| **19** | severe | 36 | change | 16 | heart | 17 |
| **20** | eye | 35 | tremor | 16 | change | 16 |
| **21** | tachycardia | 34 | severe | 16 | rashes | 16 |
| **22** | still | 33 | sleep | 14 | leg | 16 |
| **23** | month | 33 | heart rate | 14 | migraine | 15 |
| **24** | muscle | 32 | hair loss | 14 | tremor | 15 |
| **25** | sense | 32 | leg | 14 | sleep | 15 |
| **26** | time | 32 | rashes | 13 | severe | 15 |
| **27** | ear | 31 | fatigue | 13 | muscle | 14 |
| **28** | joint pain | 31 | muscle | 13 | hair loss | 14 |
| **29** | hair loss | 31 | brain fog | 13 | sensitivity | 14 |
| **30** | change | 31 | sensitivity | 13 | heart rate | 14 |
| **31** | leg | 29 | migraine | 12 | fatigue | 14 |
| **32** | tingling | 29 | tingling | 11 | ache | 13 |
| **33** | sleep | 29 | feel | 11 | time | 13 |
| **34** | migraine | 29 | light | 11 | tingling | 13 |
| **35** | long | 28 | heart | 11 | brain fog | 13 |

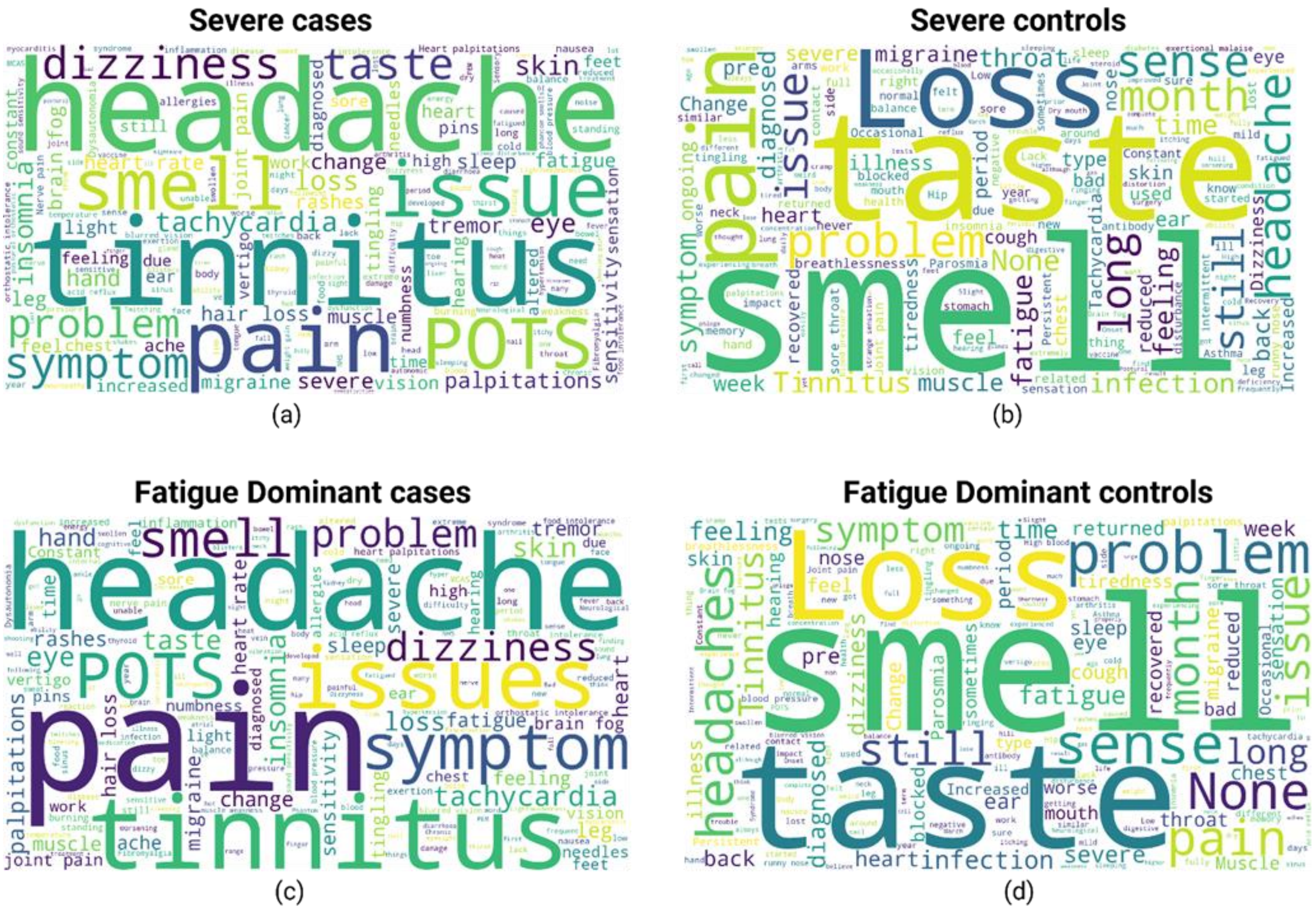

**Supplementary Figure 2:** Word clouds for (a) Severe cases, (b) Severe controls, (c) Fatigue Dominant cases and (d) Fatigue Dominant controls generated from free-text responses on other symptoms individuals in the GOLD long COVID cohort have experienced since their illness that was not directly covered in the questionnaire. The size of the words in the word clouds indicate the weights based on their incidence within each subcohort.

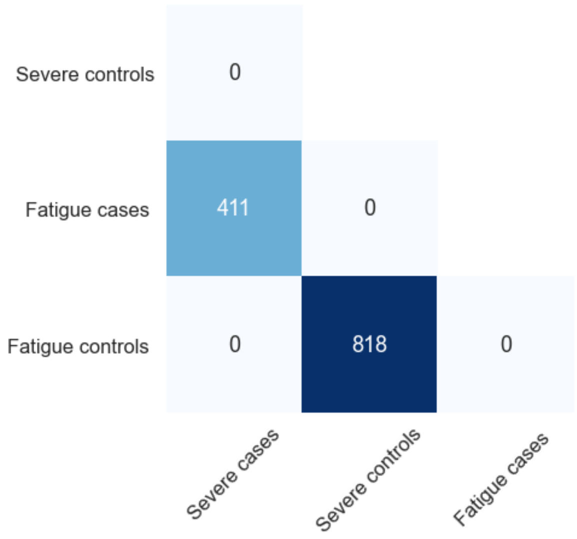

**Supplementary Figure 3:** Sample overlap between the Severe and Fatigue Dominant long COVID cohorts derived from the GOLD study.

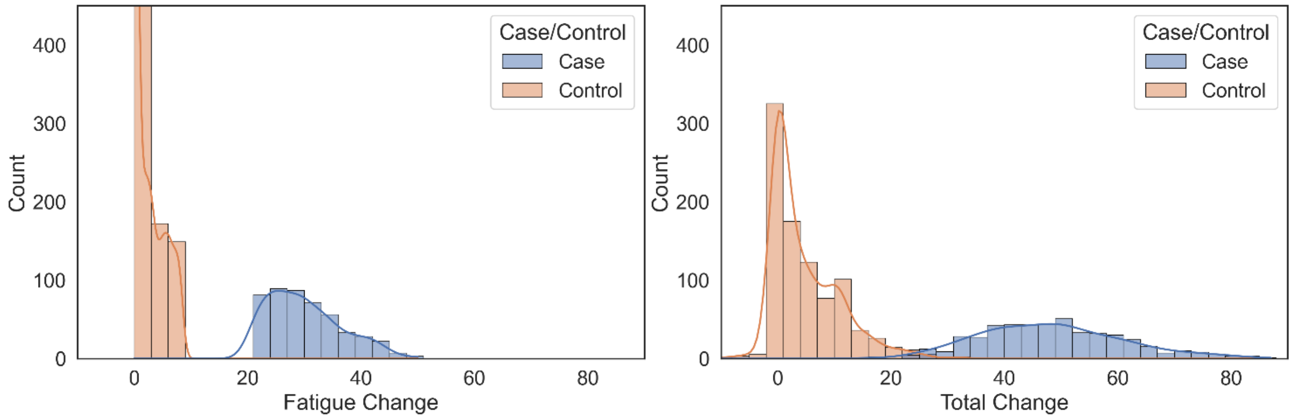

**Supplementary Figure 4:** Distribution of the 'Fatigue Change' and 'Total Change' scores for cases and controls in the Fatigue Dominant long COVID cohort.

#### 2.2 Genotype Quality Control

Appropriate quality control of genotype data was performed using GRAF[1] (Genetic Relationship and Fingerprinting) and PLINK[2] based on standard quality control procedures to ensure thorough cleaning of the data before it is used for genomic analyses.

This included the following steps:

1. Sample and SNP filtering based on missingness: The filtering for SNPs with missing data (<5%) was followed by filtering of individuals with missing data (<5%) using PLINK.
2. Minor Allele Frequency (MAF) filtering of SNPs: The genotype data would be filtered to exclude SNPs with MAF < 5% using PLINK.
3. Hardy-Weinberg Equilibrium (HWE) filtering: HWE filtering was performed on controls with $p<10^{-10}$ using PLINK.
4. Heterozygosity filtering: Samples with extreme (very high or very low) heterozygosity were removed.
5. Sample filtering based on relatedness: GRAF-rel[3] was used to identify duplicates and closely related subjects in the dataset. After identification of close relatives, only one representative of each closely related family pairs was retained.
6. Sex discrepancy of individuals: Samples that have discrepancies between the sex recorded in the dataset and their sex based on absence/presence of a Y chromosome were removed.

#### 2.3 Gene Annotation Data Sources

**Supplementary Table 3:** Example public data sources used for annotating genes in the PrecisionLife platform.

| Annotation Type | Data Source(s) |
|---|---|
| **Genomic Context** | Ensembl[4], Entrez[5], gnomAD6, HaploReg[7], ClinVar[8], Variant Effect Predictor[9], RegulomeDB[10] |
| **Encoded Protein Structure and Function** | Uniprot[11], InterPro[12], PDBe-KB[13] |
| **SNP-Disease or Trait Association** | dbSNP[14], GWAS Catalog[15], PharmGKB[16], Open Targets Genetics[17] |
| **Gene-Disease or Trait Association** | PubMed[18], PubChem[19], Open Targets[20], Mouse Genome Informatics[21], Human Phenotype Ontology[22], OMIM[23] |
| **Gene Tissue/Cell Expression** | GTEx[24], Human Protein Atlas[25], Expression Atlas[26] |

| | |
|---|---|
| **Pathways, Interactions or MoA** | Reactome[27], KEGG[28], Gene Ontology Annotation[29], PANTHER[30], WikiPathways[31], IntAct[32], STRING[33], BioGRID[34], PubMed[18] |
| **Safety / Toxicology** | International Mouse Phenotyping Consortium[35], Tox21[36], eTox[37], HeCaToS[38], Open Targets[20] |
| **Drugs and Chemical Compounds** | ChEMBL[39], DrugBank[40], PHAROS[41], PubChem[19], ProbeMiner[42], PDBe-KB[13], ClinicalTrials.gov[43], Unichem[44], OpenFDA[45] |

#### 2.4 Comparison of Long COVID with ME/CFS

**Supplementary Table 4:** List of 39 SNPs identified in disease signatures associated with long COVID in the Severe and Fatigue Dominant long COVID cohorts that can be linked to 9 genes identified in a combinatorial analysis of UK Biobank ME/CFS patients. 13 SNPs are shown in **bold** that were identified as critical SNPs in each long COVID cohort.

| Gene | Severe cohort | Fatigue-Dominant cohort |
|---|---|---|
| **AKAP1** | | rs2270278 |
| **ATP9A** | | rs1205693 |
| **ATP9A** | | rs2147338 |
| **ATP9A** | | rs6067922 |
| **ATP9A** | **rs6096573** | **rs6096573** |
| **ATP9A** | | rs6126313 |
| **ATP9A** | **rs77771672** | **rs77771672** |
| **ATP9A** | **rs2426361** | |
| **CDON** | | rs4937076 |
| **CLOCK** | | **rs62303689** |
| **GPC5** | rs1536620 | **rs1536620** |
| **GPC5** | | **rs16946160** |
| **GPC5** | rs17267214 | rs17267214 |
| **GPC5** | rs2183606 | rs2183606 |
| **GPC5** | | rs35616509 |
| **GPC5** | rs6492567 | rs6492567 |
| **GPC5** | | rs7319083 |
| **GPC5** | | rs950725 |
| **GPC5** | | rs9560999 |
| **GPC5** | | rs9589601 |
| **GPC5** | rs2149066 | |
| **GPC5** | **rs462954** | |
| **GPC5** | rs59625704 | |
| **GPC5** | **rs9301839** | |
| **GPC5** | rs9523723 | |
| **GPC5** | **rs9560843** | |
| **GPC5** | **rs989236** | |
| **INSR** | rs10427021 | rs10427021 |
| **INSR** | rs6510976 | rs6510976 |
| **INSR** | rs7317941 | rs7317941 |
| **INSR** | **rs8110533** | rs8110533 |
| **INSR** | rs10426094 | |

| | | |
|---|---|---|
| **INSR** | rs4804103 | |
| **PHACTR2** | rs7742964 | rs7742964 |
| **PHACTR2** | | rs9376793 |
| **PHACTR2** | | rs9496708 |
| **PHACTR2** | rs10979 | |
| **SLC15A4** | **rs11059915** | **rs11059915** |
| **USP6NL** | **rs11257114** | |

#### 2.5 Expanded Genotype Analysis

**Supplementary Table 5:** Descriptions of the seven categories that can potentially be assigned to critical SNPs during expanded genotype analyses.

| Category | Observed association of critical SNP minor allele with disease |
|---|---|
| Universally causative | Consistently associated with increased odds of disease relative to the homozygous wildtype genotype. |
| Universally protective | Consistently associated with decreased odds of disease relative to the homozygous wildtype genotype. |
| SNP-specific causative effect | Associated with increased odds of disease only when it co-occurs with the minor allele of at least one interacting SNP. Associated with decreased odds of disease in patients who are homozygous for the wildtype alleles of the interacting SNPs. |
| SNP-specific protective effect | Associated with decreased odds of disease only when it co-occurs with the minor allele of at least one SNP. Associated with increased odds of disease in patients who are homozygous for the wildtype alleles of the interacting SNPs. |
| Combination-specific causative effect | Associated with increased odds of disease only when it co-occurs with a specific combination of genotypes at multiple interacting SNPs. Associated with decreased odds of disease in other patients. |
| Combination-specific protective effect | Associated with decreased odds of disease only when it co-occurs with a specific combination of genotypes at multiple interacting SNPs. Associated with increased odds of disease in other patients. |
| Ambiguous | No consistent association with disease risk across expanded genotype signature comparisons. May reflect a highly complex biological interaction, a “false positive” disease signature, or stochastic noise in the data that obscures true biological relationships. |

#### 2.6 Genetic Ancestry Analysis of GOLD study

**Supplementary Table 6:** Genetic ancestry distribution of samples within the two long COVID cohorts in the GOLD study.

| | **Severe Long COVID**<br>***n* = 1,323** | | **Fatigue Dominant Long COVID**<br>***n* = 1,386** | |
|---|---|---|---|---|
| | **Cases**<br>**(*n*=459)** | **Controls**<br>**(*n*=864)** | **Cases**<br>**(*n*=477)** | **Controls**<br>**(*n*=909)** |
| **Genetic ancestry [*n*]**<br>S Asian: South Asian<br>E Asian: East Asian<br>O Asian: Other Asian<br>AA: African American | Europe: 426<br>S Asian: 14<br>E Asian: 1<br>African: 1<br>AA: 1<br>Hispanic1: 7<br>Hispanic: 0<br>O Asian: 1<br>Other: 8 | Europe: 794<br>S Asian: 23<br>E Asian: 9<br>African: 6<br>AA: 12<br>Hispanic1: 5<br>Hispanic2: 2<br>O Asian: 9<br>Other: 3 | Europe: 443<br>S Asian: 15<br>E Asian: 0<br>African: 1<br>AA: 3<br>Hispanic1: 6<br>Hispanic2: 0<br>O Asian: 1<br>Other: 8 | Europe: 838<br>S Asian: 23<br>E Asian: 10<br>African: 6<br>AA: 11<br>Hispanic1: 7<br>Hispanic2: 2<br>O Asian: 9<br>Other: 3 |

#### 2.7 PLINK GWAS Analysis

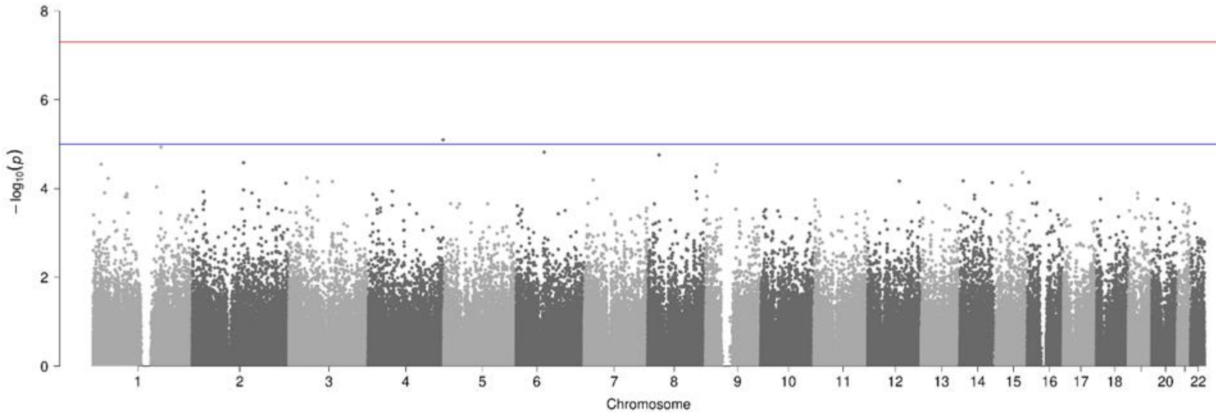

(a)

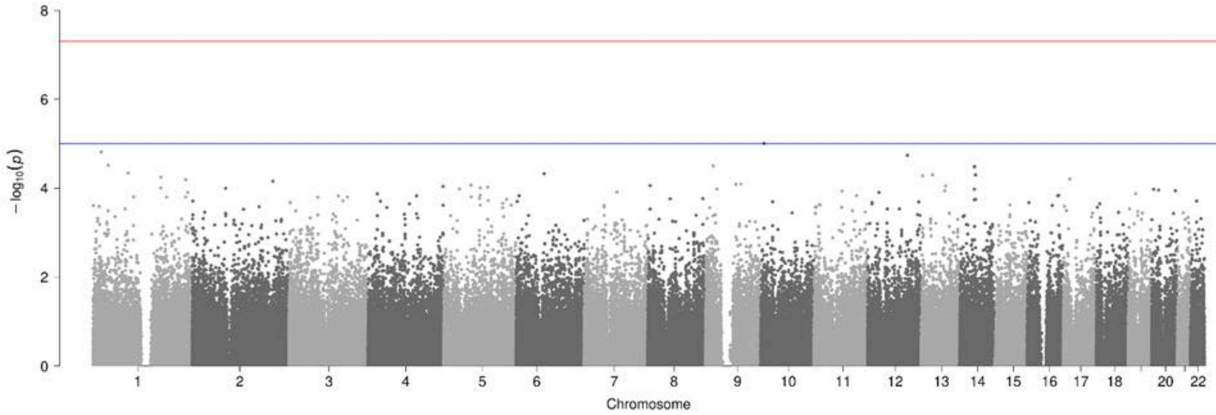

(b)

**Supplementary Figure 5:** Manhattan plots generated using PLINK of genome-wide *p*-values of association for the (a) Severe cohort (*n* = 1,323 where cases = 459 and controls = 864) and (b) Fatigue Dominant cohort (*n* = 1,386 where cases = 477 and controls = 909). The horizontal blue and red lines represent the genome-wide significance values of $p<1 \times 10^{-5}$ and $p<5 \times 10^{-8}$ respectively.

#### 2.8 Combinatorial Analysis

**Supplementary Table 7:** The number of disease signatures (SNP combinations) that pass ancestry confounder analysis and the proportion of non-European cases (based on genetic ancestry) in each long COVID cohort.

| Long COVID Cohort | Disease Signatures that pass ancestry confounder analysis | Non-European Case Count (% of non-European samples) |
|---|---|---|
| **Severe** | 1,188 (100%) | 33 (32.35%) |
| **Fatigue Dominant** | 1,306 (91.0%) | 34 (32.4%) |

#### 2.9 Tissue or Cell Expression Profile of Long COVID Genes

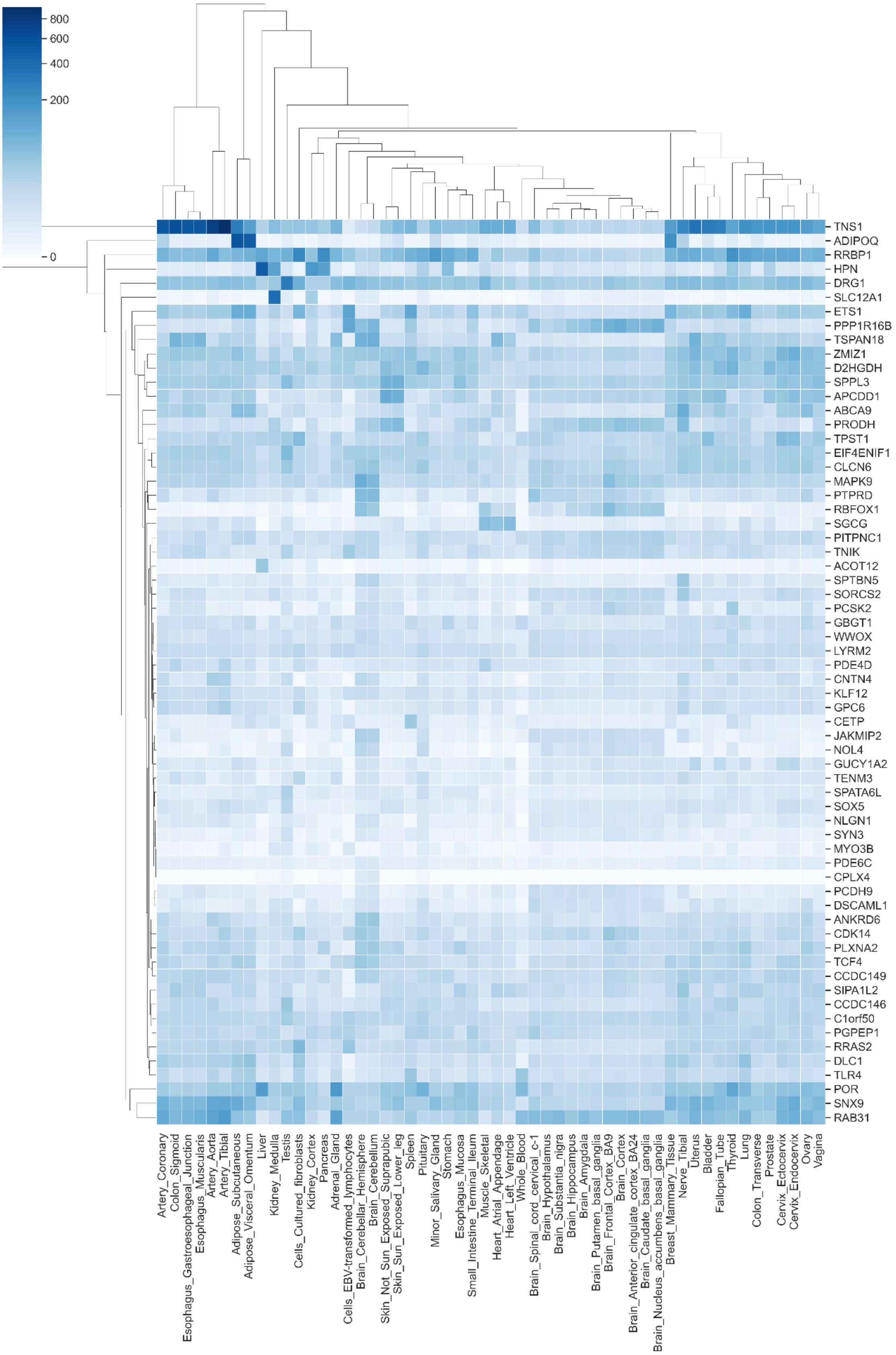

**Supplementary Figure 6:** Clustered heatmap showing tissue expression profiles from GTEx[24] for 64 out of 73 long COVID genes identified in the GOLD study with expression information.

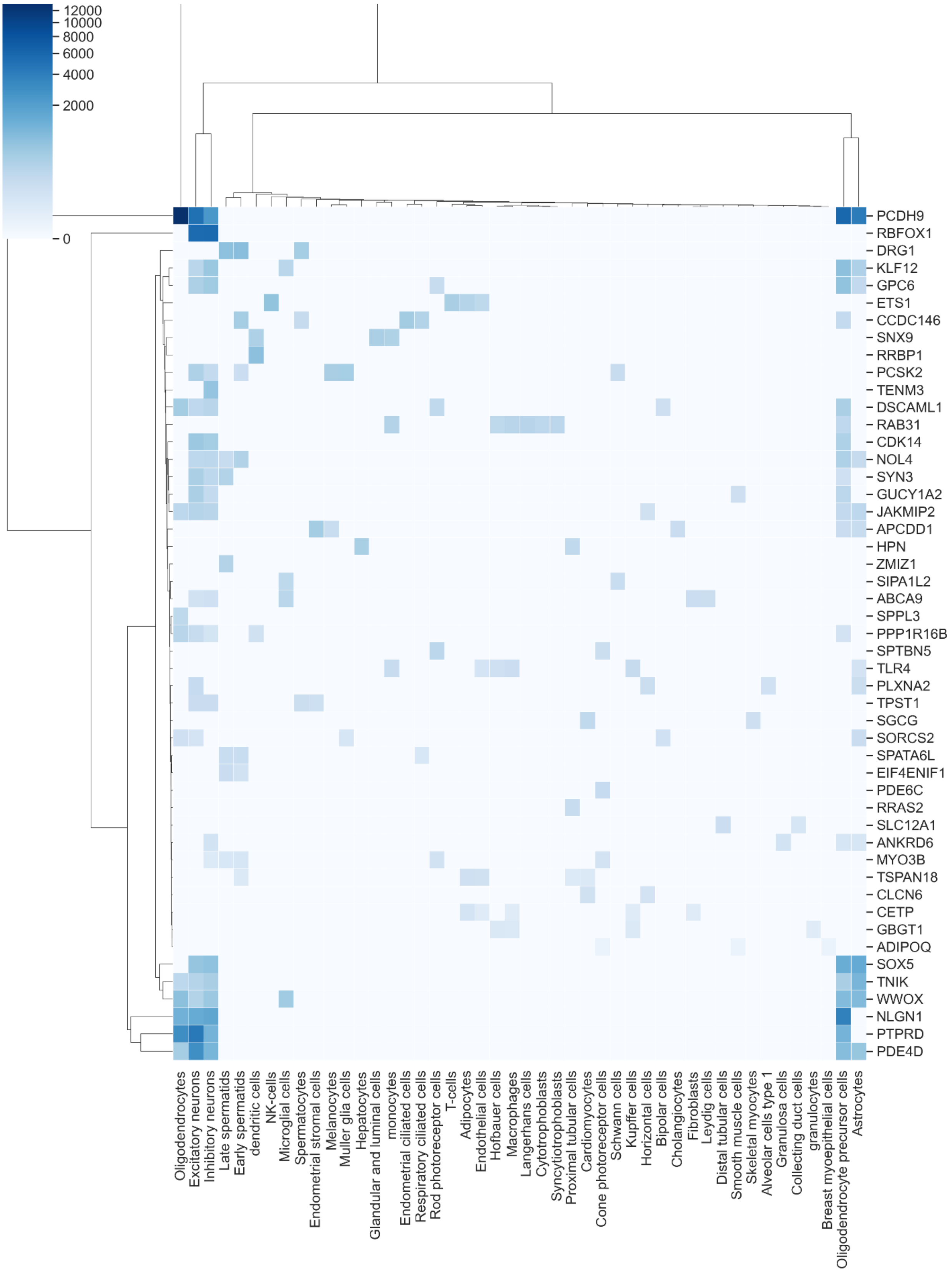

**Supplementary Figure 7:** Clustered heatmap showing single cell expression profiles from Human Protein Atlas[25] for 49 out of 73 long COVID genes identified in the GOLD study with expression information.

#### 2.10 Cross-Disease Analysis

**Supplementary Table 8:** Disease-associated genes identified for each indication group that have genetic evidence reported in Open Targets[20] (version 23.02, released in February 2023). Only genes with target-disease genetic association score > 0.9 out of 1.0 have been used.

| Indication groups | No. of disease-associated genes in indication group (Open Targets target-disease | Overlap with long COVID genes (*n* = 73) |
|---|---|---|
| **Neurodegenerative disease** (EFO:0005772) | 170 | 0 |
| **Mental or behavioral disease** (EFO:0000677) | 97 | 1 |
| **Cardiovascular disease** **(**EFO:0000319**)** | 107 | 0 |
| **Gastrointestinal disease** (EFO:0010282) | 80 | 0 |
| **Autoimmune disease** (EFO:0005140) | 40 | 1 |
| **Metabolic disease** (EFO:0000589) | 410 | 1 |

**Supplementary Table 9:** List of diseases included under each indication group based on Experimental Factor Ontology (EFO) ontology[46].

| **Indication group** (EFO term) | **Diseases included under each indication group** (direct descendants in the EFO ontology) |
|---|---|
| **Neurodegenerative disease** (EFO:0005772) | Marchiafava-Bignami Disease<br>secondary Parkinson disease<br>Primary progressive aphasia<br>tauopathy<br>motor neuron disease<br>inherited neurodegenerative disorder<br>cerebellar degeneration<br>human prion disease<br>infantile bilateral striatal necrosis<br>diffuse cerebral and cerebellar atrophy - intractab...<br>eye degenerative disorder<br>demyelinating disease<br>neuroaxonal dystrophy<br>synucleinopathy |
| **Mental or behavioral disease** (EFO:0000677) | Atypical progressive supranuclear palsy<br>Right temporal lobar atrophy<br>Classical progressive supranuclear palsy<br>CADASIL<br>pathological gambling<br>post-concussion syndrome<br>internalizing disorder<br>stress-related disorder<br>dysphoria<br>visuospatial impairment<br>somatoform disorder |

| | |
|---|---|
| | gender identity disorder<br>Sleep Disorder<br>Kluver-Bucy syndrome<br>agnosia<br>anxiety disorder<br>substance withdrawal syndrome<br>developmental disorder of mental health<br>psychosis<br>occupation-related stress disorder<br>eating disorder<br>mood disorder<br>drug dependence<br>nervousness<br>decreased attention<br>adjustment disorder<br>alcohol-induced mental disorder<br>cognitive disorder<br>drug-induced mental disorder |
| **Cardiovascular disease** (EFO:0000319) | vascular disease<br>heart disease<br>cardiovascular neoplasm<br>congenital anomaly of cardiovascular system<br>autoimmune disorder of cardiovascular system |
| **Gastrointestinal disease** (EFO:0010282) | Genetic digestive tract tumor<br>Genetic intestinal disease<br>chemotherapy-induced gastrointestinal mucositis<br>ventral hernia<br>stomach rupture<br>stomach diverticulum<br>lingual thyroid<br>immunoproliferative small intestinal disease<br>Chilaiditi Syndrome<br>enterocolitis<br>colon diverticulum<br>mouth disease<br>intestinal volvulus<br>gastric outlet obstruction<br>gastric antral vascular ectasia<br>Salivary Gland Acinic Cell Carcinoma<br>Intestinal Type Adenocarcinoma<br>Hyperplastic Polyp<br>Gastric Diffuse Large B-Cell Lymphoma<br>Duodenal Gastrin-Producing Neuroendocrine Tumor<br>Digestive System Adenoma<br>gastrointestinal toxicity<br>hepatobiliary disease<br>gastrointestinal ulceration, recurrent, with dysfunctional platelets<br>small intestine enteropathy<br>flatulence<br>stomach disease<br>pancreas disease<br>disease of peritoneum<br>foreign body in gastrointestinal tract<br>intestinal disease |

| | |
|---|---|
| | splenic disease<br>trichuriasis<br>staphyloenterotoxemia<br>paratyphoid fever<br>oesophagostomiasis<br>necatoriasis<br>hymenolepiasis<br>gastrointestinal tuberculosis<br>echinostomiasis<br>dicrocoeliasis<br>coccidiosis<br>ascaridiasis<br>ascariasis<br>rotavirus infection<br>upper digestive tract disorder<br>digestive system neuroendocrine neoplasm<br>gastrointestinal polyp<br>neoplasm of oropharynx<br>gastroesophageal disease<br>congenital enteropathy due to enteropeptidase deficiency<br>Cronkhite-Canada syndrome<br>peptic ulcer disease<br>digestive system cancer<br>diarrheal disease<br>autoimmune disorder of gastrointestinal tract<br>benign digestive system neoplasm |
| **Autoimmune disease** (EFO:0005140) | Susac Syndrome<br>oroficial granulomatosis<br>malacoplakia<br>monogenic diabetes<br>Eosinophilia-Myalgia Syndrome<br>CNS demyelinating autoimmune di...<br>STAT3 gain of function<br>latent autoimmune diabetes in a...<br>autoimmune bullous skin disease<br>reactive arthritis<br>Guillain-Barre syndrome<br>autoimmune thrombocytopenic pur...<br>autoimmune thyroid disease<br>cryoglobulinemia<br>Autoimmune Hepatitis<br>Granulomatosis with Polyangiiti...<br>Myasthenia gravis<br>anti-neutrophil antibody associ...<br>Vitiligo<br>cutaneous lupus erythematosus<br>Behcet's syndrome<br>inflammatory bowel disease<br>antiphospholipid syndrome<br>juvenile idiopathic arthritis<br>Sjogren syndrome<br>rheumatoid arthritis<br>systemic lupus erythematosus |
| **Metabolic disease** | Tumor Lysis Syndrome |

| | |
|---|---|
| (EFO:0000589) | hyperamylasemia<br>lipodystrophy<br>hemochromatosis<br>acquired metabolic diseas...<br>lactic acidosis<br>x-linked warfarin sensiti...<br>metabolic toxicity<br>mineral metabolism diseas...<br>glucose metabolism diseas...<br>methylmalonic aciduria (c...<br>hepatic methionine adenos...<br>hyperprolactinemia<br>gout<br>Hypertriglyceridemia<br>diabetic retinopathy<br>xanthoma<br>nutritional disorder<br>diabetic nephropathy<br>metabolic syndrome<br>disorder of organic acid ...<br>steroid metabolism diseas...<br>disorder of acid-base bal...<br>pyrimidine metabolism dis...<br>purine metabolism disease<br>porphyrin metabolism dise...<br>carbohydrate metabolism d...<br>hyperlipoproteinemia<br>bilirubin metabolism dise...<br>disorder of glycosylation<br>hyperlipidemia<br>proteostasis deficiencies<br>inborn errors of metaboli...<br>steroid dehydrogenase def...<br>hypoalphalipoproteinemia<br>developmental anomaly of ...<br>familial thyroid dyshormo...<br>chondrocalcinosis<br>xanthinuria<br>glutaric aciduria |

#### 2.11 Genes Previously Reported in Long COVID Transcriptomics Analysis

**Supplementary Table 10:** Table showing long COVID genes identified in the GOLD study by the PrecisionLife platform that were reported to be differentially expressed (adjusted *p* value < 0.05) by Thomas et. al[47] in different cell types (ModelVariant = 'Serology_AGM'). Upregulated genes have positive log2 fold change (logFC) and downregulated genes have negative logFC.

| Gene | Long COVID cohort | Cell Type | log2 fold change (logFC) | Adjusted *p* value |
|---|---|---|---|---|
| ***ABCA9*** | Fatigue Dominant | NK cells resting | 6.338 | 0.028 |
| ***DRG1*** | Fatigue Dominant | T cells CD8 | -0.566 | 0.025 |
| ***ETS1*** | Severe | T cells CD4 memory resting | 1.796 | 0.000 |

| | | | | |
|---|---|---|---|---|
| ***ETS1*** | Severe | T cells CD8 | -1.224 | 0.031 |
| ***ETS1*** | Severe | Neutrophils | -1.563 | 0.006 |
| ***JAKMIP2*** | Fatigue Dominant | T cells CD4 memory resting | 2.142 | 0.048 |
| ***JAKMIP2*** | Fatigue Dominant | Neutrophils | -2.330 | 0.049 |
| ***KLF12*** | Fatigue Dominant | T cells CD4 memory resting | 2.072 | 0.000 |
| ***KLF12*** | Fatigue Dominant | T cells gamma delta | -1.386 | 0.036 |
| ***KLF12*** | Fatigue Dominant | Neutrophils | -1.519 | 0.025 |
| ***LYRM2*** | Fatigue Dominant | T cells CD4 memory resting | 0.637 | 0.049 |
| ***LYRM2*** | Fatigue Dominant | T cells CD8 | -0.840 | 0.021 |
| ***LYRM2*** | Fatigue Dominant | Neutrophils | -0.842 | 0.016 |
| ***LYRM2*** | Fatigue Dominant | T cells CD8 | -1.093 | 0.002 |
| ***PPP1R16B*** | Severe | T cells CD4 memory resting | 1.785 | 0.004 |
| ***PPP1R16B*** | Severe | Neutrophils | -1.805 | 0.010 |
| ***RAB31*** | Severe | T cells CD8 | 1.054 | 0.032 |
| ***RRAS2*** | Severe | T cells CD4 memory resting | 1.730 | 0.010 |
| ***RRAS2*** | Severe | Neutrophils | -1.725 | 0.025 |
| ***RRAS2*** | Severe | T cells CD8 | -1.941 | 0.006 |
| ***SNX9*** | Severe | T cells CD4 memory resting | 1.007 | 0.028 |
| ***SPON1*** | Fatigue Dominant | Plasma cells | -7.862 | 0.021 |
| ***SPPL3*** | Fatigue Dominant | T cells CD8 | 1.120 | 0.002 |
| ***SPPL3*** | Fatigue Dominant | NK cells resting | 1.023 | 0.026 |
| ***SPPL3*** | Fatigue Dominant | T cells CD8 | 0.944 | 0.016 |
| ***SPPL3*** | Fatigue Dominant | T cells gamma delta | 0.849 | 0.040 |
| ***TCF4*** | Severe | B cells memory | 2.066 | 0.046 |
| ***TENM3*** | Severe | T cells follicular helper | 6.741 | 0.028 |

#### 2.12 Gene Tractability Analysis

**Supplementary Table 11:** Genes identified in the long COVID study with their tractability as drug targets using annotations from OpenTargets[20] and other annotations from Human Protein Atlas[25] and PHAROS[41].

| Gene | Long COVID Cohort | Protein class | PHAROS target classification | Small molecule Tractability | Small molecule Clinical | Antibody Tractability | Drugs | Active Compounds |
|---|---|---|---|---|---|---|---|---|
| ***GUCY1A2*** | Severe/Fatigue | Enzyme | Tclin | 0.3 | 1 | 0 | 8 | 595 |
| ***CETP*** | Severe/Fatigue | Transporter | Tchem | 1 | 0.7 | 1 | 4 | 555 |
| ***TLR4*** | Severe | Enzyme, | Tchem | 0.3 | 0.7 | 1 | 10 | 506 |

| | | | | | | | | |
|---|---|---|---|---|---|---|---|---|
| | | Transporter | | | | | | |
| ***TCF4*** | Severe/Fatigue | Transcription factor | Tbio | 0 | 0 | 0 | 0 | 334 |
| ***MYO3B*** | Severe/Fatigue | Enzyme | Tchem | 0.3 | 0 | 0 | 0 | 245 |
| ***LARGE1*** | Severe/Fatigue | | Tbio | 0 | 0 | 0 | 0 | 99 |
| ***PCSK2*** | Severe/Fatigue | Enzyme | Tchem | 0 | 0 | 0.7 | 2 | 8 |
| ***PDE6C*** | Severe/Fatigue | Enzyme | Tclin | 0.3 | 1 | 0.7 | 2 | 1 |
| ***CLCN6*** | Severe/Fatigue | | Tchem | 0 | 0 | 0.7 | 0 | 2 |
| ***SOX5*** | Severe/Fatigue | Transcription factor | Tbio | 0 | 0 | 0 | 0 | 2 |
| ***PRODH*** | Severe/Fatigue | Enzyme | Tbio | 0 | 0 | 0 | 2 | 0 |
| ***RRAS2*** | Severe/Fatigue | RAS pathway related protein | Tbio | 0.7 | 0 | 0.7 | 0 | 0 |
| ***ADIPOQ*** | Severe | Transporter | Tbio | 0 | 0 | 1 | 0 | 0 |
| ***ADGRA3*** | Severe/Fatigue | G-protein coupled receptor | Tbio | 0 | 0 | 0 | 0 | 1 |
| ***SNX9*** | Severe/Fatigue | | Tbio | 0.7 | 0 | 0.3 | 0 | 0 |
| ***PPP1R16B*** | Severe/Fatigue | | Tbio | 0 | 0 | 1 | 0 | 0 |
| ***DSCAML1*** | Severe | | Tbio | 0 | 0 | 1 | 0 | 0 |
| ***SORCS2*** | Severe/Fatigue | | Tbio | 0 | 0 | 0.7 | 0 | 0 |
| ***APCDD1*** | Severe/Fatigue | | Tbio | 0 | 0 | 0.7 | 0 | 0 |
| ***ETS1*** | Severe | Transcription factor | Tbio | 0.7 | 0 | 0 | 0 | 0 |
| ***SGCG*** | Severe/Fatigue | | Tbio | 0 | 0 | 0.3 | 0 | 0 |
| ***GPC6*** | Severe/Fatigue | | Tbio | 0 | 0 | 0.3 | 0 | 0 |
| ***RAB31*** | Severe/Fatigue | | Tbio | 0 | 0 | 0.3 | 0 | 0 |
| ***MAPK9*** | Fatigue/Severe | Enzyme | Tchem | 1 | 0.7 | 0 | 6 | 877 |
| ***TNIK*** | Fatigue/Severe | Enzyme | Tchem | 1 | 0 | 0 | 1 | 765 |
| ***CDK14*** | Fatigue/Severe | Enzyme | Tchem | 0.3 | 0.7 | 0.3 | 0 | 258 |
| ***HPN*** | Fatigue/Severe | Enzyme | Tchem | 0 | 0 | 1 | 4 | 179 |
| ***POR*** | Fatigue | Enzyme | Tbio | 0.3 | 0 | 0 | 13 | 30 |
| ***PDE4D*** | Fatigue/Severe | Enzyme | Tclin | 1 | 1 | 1 | 26 | 4 |
| ***SLC12A1*** | Fatigue/Severe | Transporter | Tclin | 0.3 | 1 | 0.3 | 19 | 0 |
| ***DRG1*** | Fatigue/Severe | | Tbio | 0 | 0 | 0 | 0 | 4 |
| ***NLGN1*** | Fatigue/Severe | Transporter | Tbio | 0.3 | 0 | 1 | 0 | 0 |
| ***RRBP1*** | Fatigue | | Tbio | 0 | 0 | 0 | 1 | 0 |
| ***TNS1*** | Fatigue | | Tbio | 0 | 0 | 0.7 | 0 | 0 |
| ***ACOT12*** | Fatigue | Enzyme, Transporter | Tbio | 0.7 | 0 | 0 | 0 | 0 |
| ***SPON1*** | Fatigue/Severe | | Tbio | 0.7 | 0 | 0 | 0 | 0 |
| ***SYN3*** | Fatigue/Severe | | Tbio | 0.7 | 0 | 0 | 0 | 0 |
| ***PLXNA2*** | Fatigue/Severe | | Tbio | 0 | 0 | 0.3 | 0 | 0 |
| ***CNTN4*** | Fatigue/Severe | | Tbio | 0 | 0 | 0.3 | 0 | 0 |
| ***PTPRD*** | Fatigue/Severe | Enzyme | Tbio | 0 | 0 | 0.3 | 0 | 0 |

| | | | | | | | | |
|---|---|---|---|---|---|---|---|---|
| ***SPPL3*** | Fatigue/Severe | Enzyme | Tbio | 0 | 0 | 0.3 | 0 | 0 |
| ***WWOX*** | Fatigue/Severe | | Tbio | 0 | 0 | 0.3 | 0 | 0 |

#### 2.13 Gene Disease Associations

**Supplementary Table 12:** Genes with prior association to type 2 diabetes in OpenTargets[Error! Reference source not found.] database.

| Long COVID Cohort | Gene | Known association to type 2 Diabetes (OpenTargets) |
|---|---|---|
| Severe | ***ADIPOQ*** | Scientific literature, Animal models |
| Severe | ***ZMIZ1*** | Genetic association |
| Severe | ***PDE6C*** | Drugs approved and/or in clinical development for indication |
| Severe | ***SGCG*** | Genetic association |
| Severe | ***RAB31*** | Genetic association |
| Severe | ***NOL4*** | Genetic association |
| Severe | ***ETS1*** | Genetic association |
| Fatigue Dominant | ***TNS1*** | Genetic association |
| Fatigue Dominant | ***NLGN1*** | Genetic association |
| Fatigue Dominant | ***PITPNC1*** | Genetic association |
| Fatigue Dominant | ***EIF4ENIF1*** | Genetic association |

**Supplementary Table 13:** 19 pathways that are significantly enriched in the 73 long COVID genes that were also significantly enriched in at least one of the following indication groups - neurodegenerative, mental or behavioral, cardiovascular, gastrointestinal, autoimmune and metabolic disorders.

| GO Biological process | Long COVID (*p* value) | Autoimmune disease (*p* value) | Cardiovascular disease (*p* value) | Gastrointestinal disease (*p* value) | Mental or behavioural disease (*p* value) | Metabolic disease (*p* value) | Neurodegenerative disease (*p* value) |
|---|---|---|---|---|---|---|---|
| **anatomical structure development** | 0.022 | 0.001673 | 1.01E-14 | 0.00237 | 3.13E-17 | 6.84E-06 | 1.05E-09 |
| cell adhesion | 0.026 | 2.60E-05 | 0.000181 | | | | |
| **cell communication** | 0.039 | 1.76E-06 | 8.95E-07 | 0.032485 | 0.000232 | | 0.009076 |
| cell differentiation | 0.008 | 0.000395 | 1.39E-09 | 0.004574 | 1.71E-13 | | 3.31E-06 |
| cell-cell signaling | 0.021 | 0.594388 | 9.09E-05 | 0.001775 | 1.77E-06 | | 0.000316 |
| chemical synaptic transmission | 0.048 | | 0.037426 | | 1.03E-06 | | 3.88E-06 |
| dendrite self-avoidance | 0.045 | | | | | | |
| **filopodium assembly** | 0.045 | | | | | | |
| **foam cell differentiation** | 0.014 | | | | | 0.02689 | |
| **glial cell migration** | 0.033 | | | | 0.001138 | | |
| **modulation of chemical synaptic transmission** | 0.008 | | 0.003983 | | 2.40E-08 | | 0.004787 |
| nervous system development | 0.008 | | 5.16E-05 | 0.023153 | 2.17E-22 | 9.30E-06 | 1.88E-18 |
| neurogenesis | 0.008 | | 0.000267 | 0.036673 | 7.46E-18 | | 2.72E-12 |
| neuron cell-cell adhesion | 0.042 | | | | | | |
| **positive regulation of cell differentiation** | 0.034 | 0.000192 | 0.000622 | 0.0085 | 1.75E-05 | | |
| **positive regulation of hormone metabolic process** | 0.039 | | | | | | |
| **positive regulation of protein dephosphorylatio n** | 0.027 | | | | | | |
| **presynapse assembly** | 0.028 | | | | | | |
| **regulation of biological quality** | 0.004 | | 6.66E-12 | 0.000111 | 1.41E-05 | 1.41E-06 | 0.000444 |
| regulation of cell communication | 0.006 | 2.31E-05 | 3.25E-06 | 6.50E-05 | 1.24E-05 | | |
| **regulation of cellular component biogenesis** | 0.028 | | 0.000692 | | 0.007012 | | 0.000388 |
| regulation of signaling | 0.006 | 2.28E-05 | 2.07E-05 | 6.17E-05 | 1.18E-05 | | |
| **regulation of trans-synaptic signaling** | 0.008 | | 0.004034 | | 2.44E-08 | | 0.004861 |
| response to epinephrine | 0.039 | | 0.010242 | | | | |
| **response to stimulus** | 0.029 | 2.53E-06 | 3.65E-08 | 0.04138 | 8.65E-07 | 0.011387 | 0.01398 |
| synaptic membrane adhesion | 0.01 | | | | | | |

| | |
|---|---|
| **synaptic vesicle clustering** | 0.045 |
